# Estimates of global SARS-CoV-2 infection exposure, infection morbidity, and infection mortality rates

**DOI:** 10.1101/2021.01.24.21250396

**Authors:** Houssein H. Ayoub, Ghina R. Mumtaz, Shaheen Seedat, Monia Makhoul, Hiam Chemaitelly, Laith J. Abu-Raddad

## Abstract

We aimed to estimate, albeit crudely and provisionally, national, regional, and global proportions of respective populations that have been infected with SARS-CoV-2, and to assess infection morbidity and mortality rates, factoring both documented and undocumented infections. The estimates were generated by applying mathematical models to 159 countries and territories. The percentage of the world’s population that has been infected as of 31 December 2020 was estimated at 12.56% (95% CI: 11.17-14.05%). It was lowest in the Western Pacific Region at 0.66% (95% CI: 0.59-0.75%) and highest in the Americas at 41.92% (95% CI: 37.95-46.09%). The global infection fatality rate was 10.73 (95% CI: 10.21-11.29) per 10,000 infections. Globally per 1,000 infections, the infection acute-care bed hospitalization rate was 19.22 (95% CI: 18.73-19.51), the infection ICU bed hospitalization rate was 4.14 (95% CI: 4.10-4.18), the infection severity rate was 6.27 (95% CI: 6.18-6.37), and the infection criticality rate was 2.26 (95% CI: 2.24-2.28). If left unchecked with no interventions, the pandemic would eventually cause 8.18 million (95% CI: 7.30-9.18) deaths, 163.67 million (95% CI: 148.12-179.51) acute-care hospitalizations, 33.01 million (95% CI: 30.52-35.70) ICU hospitalizations, 50.23 million (95% CI: 46.24-54.67) severe cases, and 17.62 million (95% CI: 16.36-18.97) critical cases. The global population remains far below the herd immunity threshold and at risk of repeated waves of infection. Global epidemiology reveals immense regional variation in infection exposure and morbidity and mortality rates.

## Introduction

The severe acute respiratory syndrome coronavirus 2 (SARS-CoV-2) pandemic continues to be a global health challenge with profound adverse consequences for human health, societies, and economies [1]. While our understanding of the epidemiology of SARS-CoV-2 infection and its Coronavirus Disease 2019 (COVID-19) disease burden has progressed in the year since it emerged, two questions remain largely unanswered.

1. “To what extent have populations of individual countries and the global population been infected by this virus, regardless of whether those infections have been documented?” Infections include documented cases involving laboratory-confirmed diagnosis and undocumented asymptomatic or mild cases. While a growing number of serological surveys are being conducted to answer this question [2-10], the scope, scale, and geographic coverage of such studies remain limited.
2. “What are the *true* national and global COVID-19 morbidity and mortality rates, that is factoring all documented and undocumented infections?”

Recent scientific developments furnish an opportunity to provide answers, albeit crude approximations, to these questions. The growing number of serological surveys and analyses of national databases for this infection have shown that only about one in every ten infections have actually been diagnosed [2-11]. Moreover, a recent comprehensive analysis assessed the true infection morbidity and mortality rates for each age group, factoring both documented and undocumented infections [12].

Building on these developments, the objective of this study was to provide key provisional epidemiologic estimates nationally, regionally, and globally. These include estimates of the proportion of each population that has been already infected, estimates for the average incidence rate of this infection since epidemic onset, and estimates for overall (total population) infection acute-care and intensive-care-unit (ICU) hospitalization rates, infection severity and criticality rates, and infection fatality rate.

## METHODS

### Definitions of epidemiologic outcome measures

Two criteria for classifying infection morbidity were used: one based on hospital admissions (acute-care or ICU) and one based on clinical presentations, as per the World Health Organization (WHO) classifications of disease severity (Table 1) [13]. While the two measures overlap, with severe cases typically admitted to acute-care beds and critical cases admitted to ICU beds, mild or moderately ill COVID-19 cases are sometimes hospitalized out of caution, because of other, concurrent indications, or as a form of isolation [12].

**Table 1.** Epidemiologic outcome measures estimated in this study for each country, by region, and globally.

| Outcome measure | Definition | Interpretation |
| --- | --- | --- |
| **Infection morbidity rates** |  |  |
| **Morbidity based on hospitalization type** |  |  |
| 1. Infection acute-care bed hospitalization rate | Cumulative number of hospital admissions into acute-care beds over the cumulative number of infections, *documented and undocumented* | Proportion of infections that progress to acute-care bed hospital admission. |
| 2. Infection ICU bed hospitalization rate | Cumulative number of hospital admissions into ICU beds over the cumulative number of infections, *documented and undocumented* | Proportion of infections that progress to ICU bed hospital admission. |
| 3. Infection *total* hospitalization rate (measures 1 and 2 combined) | Cumulative number of hospital admissions into both acute-care and ICU beds over the cumulative number of infections, *documented and undocumented* | Proportion of infections that progress to acute-care or ICU bed hospital admission. |
| **Morbidity based on clinical presentation\*** |  |  |
| 4. Infection severity rate | Cumulative number of COVID-19 severe infections* over the cumulative number of infections, *documented and undocumented* | Proportion of infections that progress to become severe infections. |
| 5. Infection criticality rate | Cumulative number of COVID-19 critical infections* over the cumulative number of infections, *documented and undocumented* | Proportion of infections that progress to become critical infections. |
| 6. Infection *total* severity rate (measures 4 and 5 combined) | Cumulative number of both COVID-19 severe or critical infections* over the cumulative number of infections, *documented and undocumented* | Proportion of infections that progress to become severe or critical infections. |
| **Infection mortality rate** |  |  |
| 1. Infection fatality rate | Cumulative number of COVID-19 deaths over the cumulative number of infections, *documented and undocumented* | Proportion of infections that end in COVID-19 death. |
| **Infection occurrence metrics** |  |  |
| 1. Proportion of the population infected | Cumulative number of infections, *documented and undocumented*, over the total population size | Proportion of the population that has already infected sometime after the virus' introduction into the population. |
| 2. Infection incidence rate (also known as force of infection or hazard rate of infection) | Number of new infections *documented and undocumented* per unit time (here week) over the size of the population that is still susceptible to the infection | Rate of new infections per person-weeks. |

\*Per World Health Organization infection severity classification [13].

Three types of outcomes were estimated nationally, regionally, and globally (Table 1). The first two include *infection morbidity and mortality rates*, calculated as the cumulative number of a disease outcome (such as COVID-19 hospitalization or death) over the estimated cumulative number of infections, *documented and undocumented*. The third type includes two kinds of infection occurrence metrics, the proportion of the population that has been already infected and the infection incidence rate, both as of December 31, 2020.

### Estimations of epidemiologic outcome measures

#### Infection morbidity and mortality rates

The overall (total population) infection acute-care and ICU hospitalization rates, infection severity and criticality rates, and infection fatality rate were estimated for each country by applying the recently estimated *age-stratified* rates for these outcomes [12] to the population age-structure of each country. Age-stratified rates were based on a detailed analysis of the epidemic in Qatar [12] using data from a series of serological surveys [8-10] and extensive time-series and age-stratified data for PCR laboratory-confirmed infections, PCR testing positivity rate, antibody testing positivity rate, PCR surveys, daily hospital admissions in acute-care and ICU beds, hospital occupancy in acute-care and ICU beds, incidence of severe and critical infections, as per WHO classifications [13], and COVID-19 deaths as per WHO guidelines [14]. Qatar has one of the world’s most extensive databases to document this epidemic and its toll at the national level [15], such that Qatar’s epidemic has been one of the most thoroughly investigated and best characterized [8-12, 15-21].

It is presently unknown whether the infection morbidity and mortality rates for *each age group* (not for the total population) vary considerably from one country to another. These rates probably reflect primarily the basic biology of this infection more than the COVID-19 response or other factors in each country or population. The aim of the present study is to provide crude estimates for these rates in the *total* population of each country by factoring the population age structure, given the prominent role of age in the epidemiology of this pandemic [22-25], while future studies investigate and elaborate possible variations in these rates across different countries and populations.

Infection morbidity and mortality rates were estimated for each country and territory with a population size >1 million, as of 2020. In total, estimates were generated for 159 countries and territories, virtually covering the world population [26]. Population sizes and demographic age-structures were extracted from the United Nations World Population Prospects database [26]. In each setting, rates were derived by weighting each rate in each age group by the proportion of the population in that age group, and then summing the contributions of all age groups.

#### Incidence rate and proportion of the population infected

Two methods were used to derive the proportion of the population infected in each country and the average incidence rate since onset of the epidemic as of the end of 2020. The final estimate for each of these measures was based on the average of the estimates of both methods, to minimize the effect of potential bias inherent in each method.

### Reported COVID-19 deaths method

The first method was based on the reported number of COVID-19 deaths in each country, as per the WHO COVID-19 Dashboard [27]. The proportion of the population infected, irrespective of whether the infection was documented or undocumented, was estimated using the following expressions:

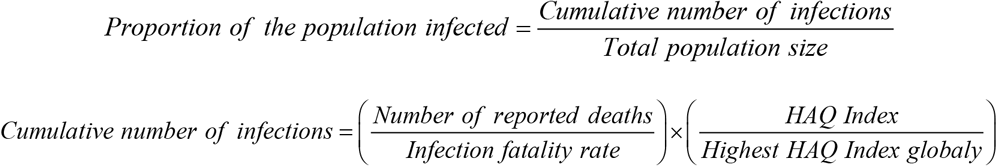

Since COVID-19 mortality may be affected by access to and quality of healthcare, with higher mortality for inferior access and quality, this method adjusts for these factors by utilizing the Global Burden of Disease study’s Healthcare Access and Quality (HAQ) Index for each country and territory [28]. The HAQ Index provides a score ranging between 0–100 [28].

The above expressions still require adjustment for the average time delay between *onset of infection* and *COVID-19 death*, estimated from studies in different countries at about 20 days [29-32]. That adjustment was incorporated by assuming that the above estimated proportion of the population infected occurred *20 days earlier* than the current time *t*. Then the *average* incidence rate of infection (*λ*), from the onset of the epidemic (at time *t0*) until the present time (time *t*), was derived using the expression:

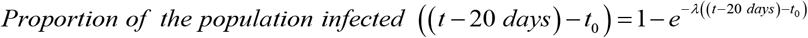

The time *t0* was set as the day of the first reported COVID-19 case in each country [27]. The derived incidence rate was then used to estimate the proportion of the population infected at the time of this study, by applying the same expression, but at time *t* (that is at December 31, 2020), instead of *t* − 20 *days*.

### Reported COVID-19 cases method

The second method was based on the reported cumulative number of laboratory-confirmed SARS-CoV-2 infections as of December 31, 2020, as reported in the WHO COVID-19 Dashboard [27]. The cumulative number of infections, *documented and undocumented*, was estimated using the following expression:

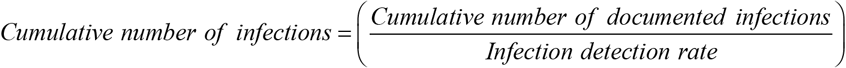

The infection detection rate is defined as the *proportion of all infections that were actually diagnosed and laboratory-confirmed*. Serological surveys and extensive analyses have shown that only about one in every ten actual infections is ever diagnosed [2-10]. Given the quality estimate for the infection detection rate in the well-characterized epidemic of Qatar, based on a series of serological surveys [8-10] and analyses of national databases, a value of 11.1% (95% uncertainty interval: 10.8-11.3%) was assumed for the infection detection rate [11, 12].

However, to account for variation in the quality of SARS-CoV-2 testing across countries, this estimate was adjusted using the HAQ Index [28]:

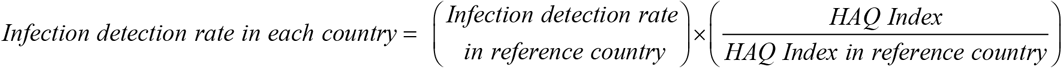

With the above expressions, and Qatar as the reference country, a second estimate was generated for each country of the proportion of the total population infected and the average incidence rate since epidemic onset.

### Uncertainty analysis

The 95% credible interval (CI) for each estimated epidemiologic outcome measure was derived by factoring the uncertainty interval of each variable used in the above equations and combining the uncertainties so as to generate the widest credible interval for each estimate.

### Reporting of estimates

The various estimated epidemiologic outcome measures and the 95% CI were reported by country or territory, regionally by WHO region, and globally. The WHO regions include the African Region (AFRO), the Region of the Americas (AMRO), the Eastern Mediterranean Region (EMRO), the European Region (EURO), the South-East Asia Region (SEARO), and the Western Pacific Region (WPRO) [33] (Figure 1).

**Figure 1:**
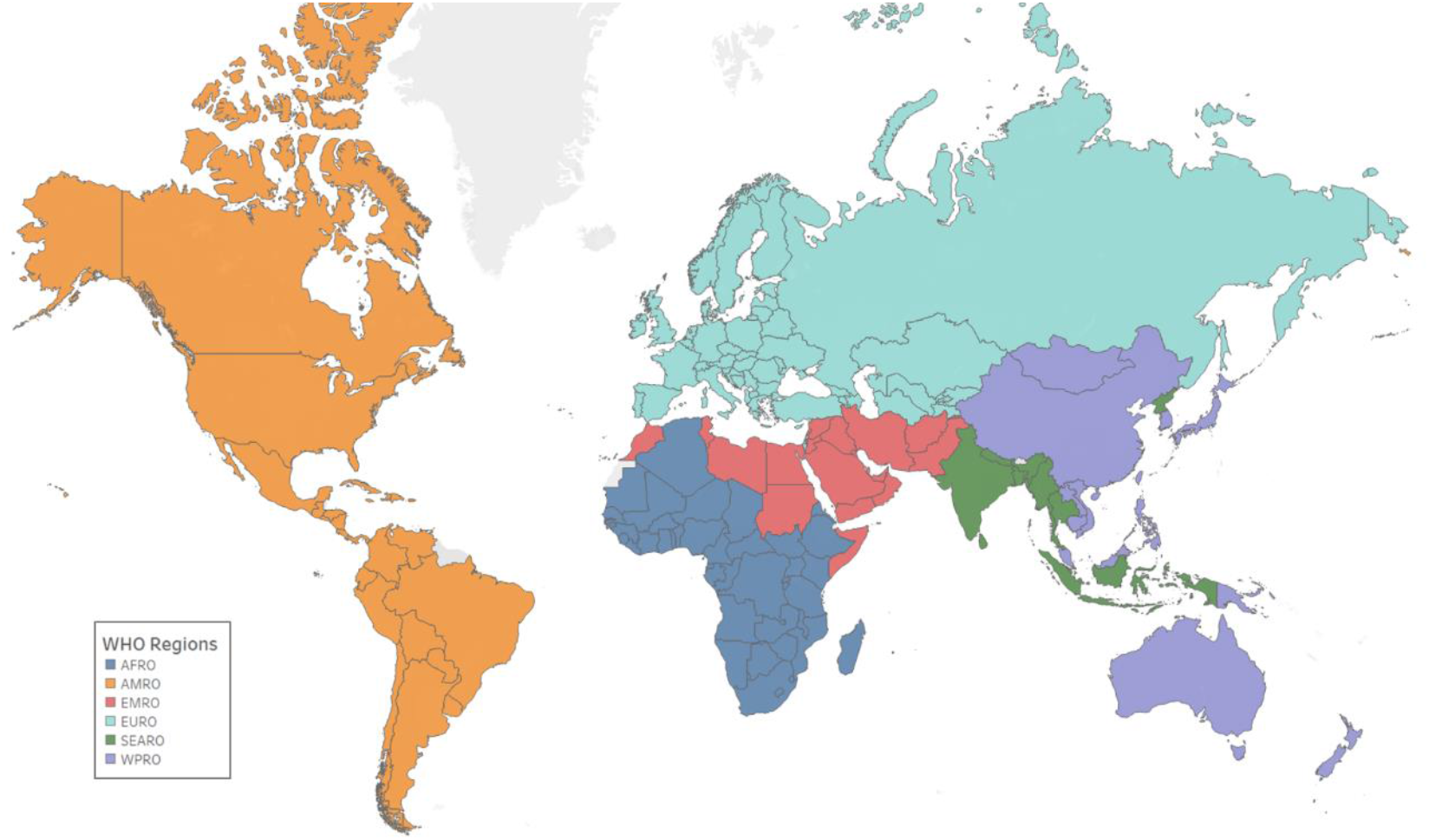
A global map of the World Health Organization (WHO) regions. The WHO regions include the African Region (AFRO) (blue), the Region of the Americas (AMRO) (orange), the Eastern Mediterranean Region (EMRO) (red), the European Region (EURO) (cyan), the South-East Asia Region (SEARO) (purple), and the Western Pacific Region (WPRO) (green) [33]. The map was generated using Tableau 10.1 software [45].

Mathematical modeling analyses were conducted in MATLAB R2019a (Boston/MA/USA) [34].

## RESULTS

The various estimated outcome measures for each country and territory are listed in Tables S1-S6 in the Supporting Information. An overview of results by WHO region and globally is provided below.

The estimated infection acute-care bed hospitalization rate, infection ICU bed hospitalization rate, infection severity rate, and infection criticality rate were lowest in AFRO and highest in EURO, with substantial variation across regions (Figure 2). Globally, the infection acute-care bed hospitalization rate was 19.22 (95% CI: 18.73-19.51) per 1,000 infections, the infection ICU bed hospitalization rate was 4.14 (95% CI: 4.10-4.18) per 1,000 infections, the infection severity rate was 6.27 (95% CI: 6.18-6.37) per 1,000 infections, and the infection criticality rate was 2.26 (95% CI: 2.24-2.28) per 1,000 infections. Meanwhile, the infection total hospitalization rate was 23.36 (95% CI: 22.83-23.69) per 1,000 infections and the infection total severity rate was 8.53 (95% CI: 8.42-8.65) per 1,000 infections (Figure 2).

**Figure 2:**
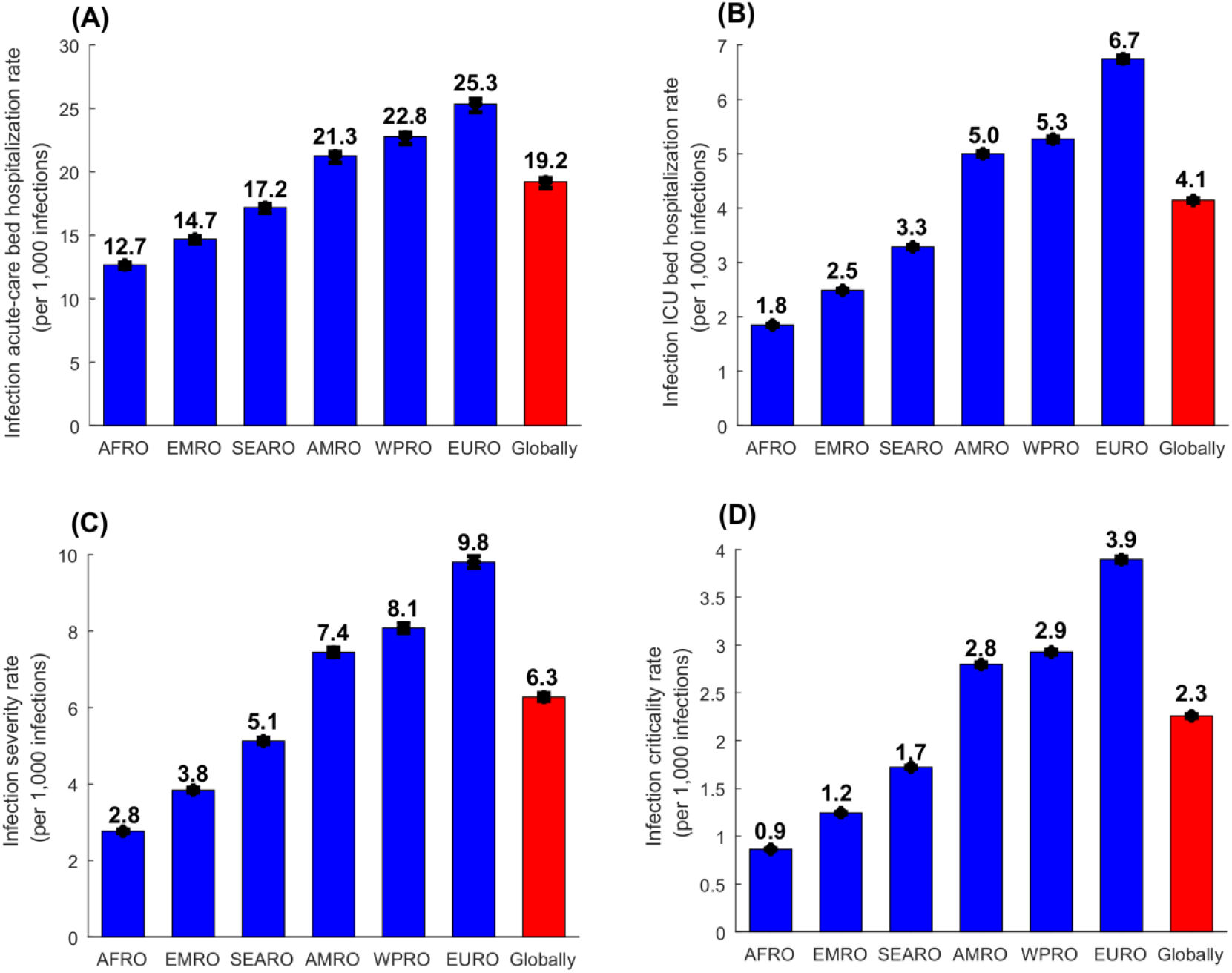
Estimated A) infection acute-care bed hospitalization rates, B) infection ICU bed hospitalization rates, C) infection severity rates, and D) infection criticality rates, across WHO regions and globally. WHO regions include the African Region (AFRO), the Eastern Mediterranean Region (EMRO), the South-East Asia Region (SEARO), the Region of the Americas (AMRO), the Western Pacific Region (WPRO), and the European Region (EURO) (Figure 1). Classification of infection severity and criticality was per WHO infection severity criteria [13].

The estimated infection fatality rate per 10,000 infections, was lowest in AFRO at 3.57 (95% CI: 3.39-3.76), followed by EMRO at 5.40 (95% CI: 5.09-5.64), SEARO at 7.64 (95% CI: 7.26-8.05), AMRO at 13.80 (95% CI: 13.13-14.51), WPRO at 13.92 (95% CI: 13.24-14.65), and highest in EURO at 20.19 (95% CI: 19.23-21.21) (Figure 3). Globally, the infection fatality rate was 10.73 (95% CI: 10.21-11.29) per 10,000 infections.

**Figure 3:**
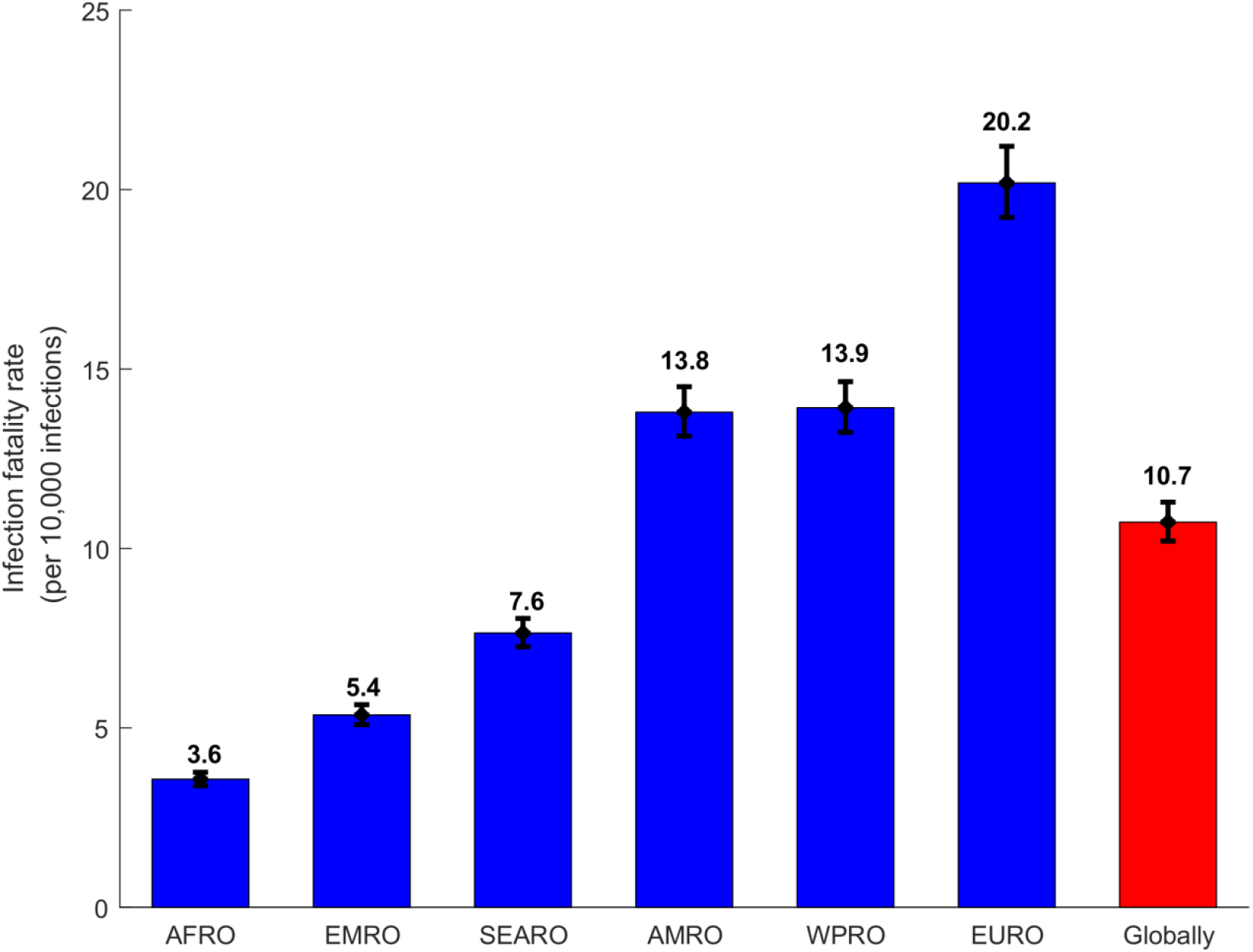
Estimated infection fatality rates, across WHO regions and globally. WHO regions include the African Region (AFRO), the Eastern Mediterranean Region (EMRO), the South-East Asia Region (SEARO), the Region of the Americas (AMRO), the Western Pacific Region (WPRO), and the European Region (EURO) (Figure 1). Classification of COVID-19 mortality was per WHO criteria [14].

The estimated incidence rate, across WHO regions and globally, using the reported deaths method was higher than that using the reported cases method, for all regions other than SEARO (Figure 4A). The averaged incidence rate, per 10,000 person-weeks, was lowest in WPRO at 1.4 (95% CI: 1.2-1.6), followed by AFRO at 8.7 (95% CI: 7.2-10.5), SEARO at 18.1 (95% CI: 15.6-20.8), EMRO at 40.5 (95% CI: 33.4-49.7), EURO at 67.4 (95% CI: 58.8-77.1), and highest in AMRO at 131.7 (95% CI: 113.6-154.2) (Figure 4B). Globally, the averaged incidence rate was 35.4 (95% CI: 30.5-41.4) per 10,000 person-weeks.

**Figure 4:**
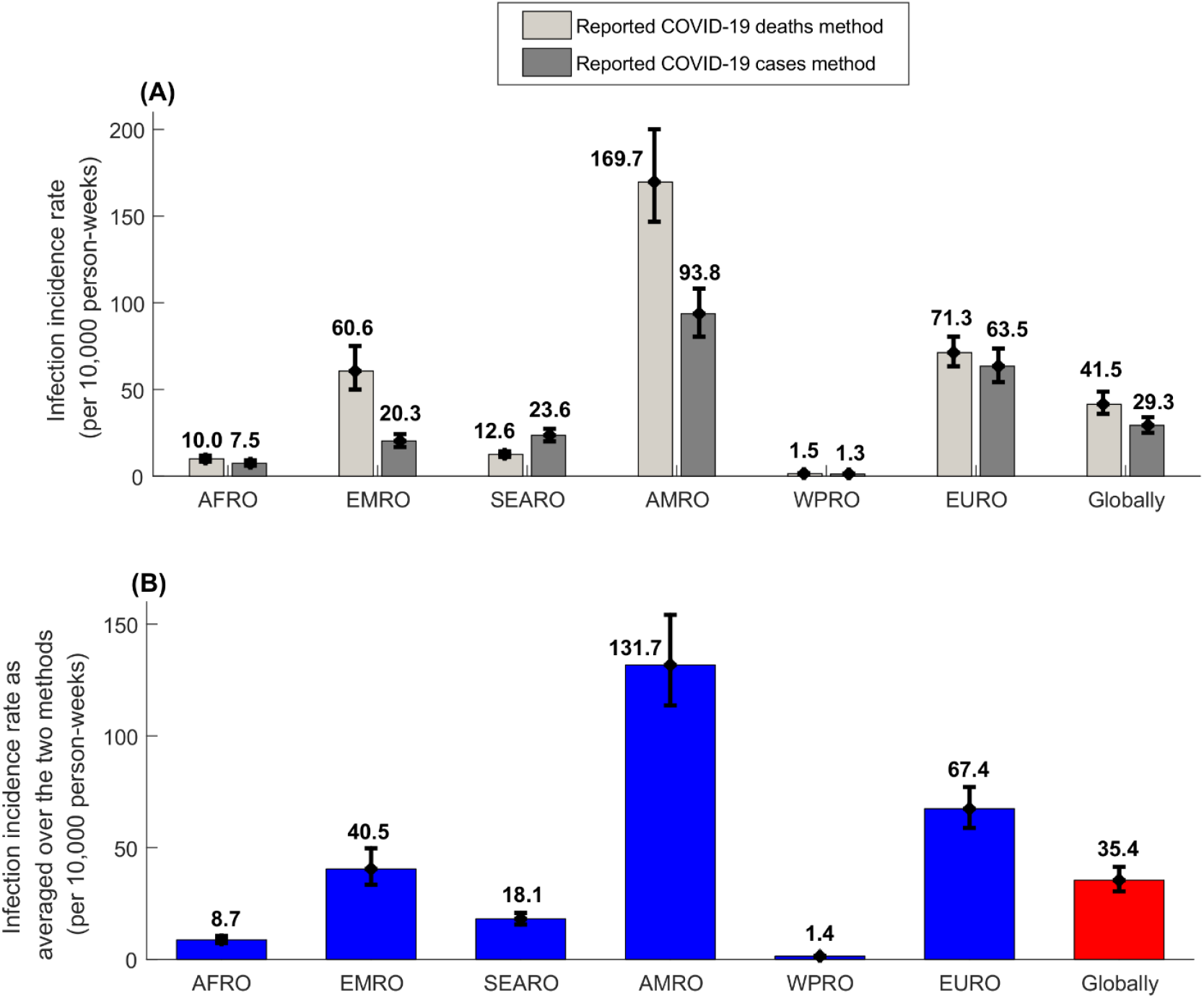
Estimated infection incidence rates across WHO regions and globally. A) Estimated average infection incidence rates since epidemic onset using the reported COVID-19 deaths method and using the reported COVID-19 cases method. B) Estimated infection incidence rates as averaged over the two methods. WHO regions include the African Region (AFRO), the Eastern Mediterranean Region (EMRO), the South-East Asia Region (SEARO), the Region of the Americas (AMRO), the Western Pacific Region (WPRO), and the European Region (EURO) (Figure 1).

The estimated percentage of the population infected by the end of 2020, across WHO regions and globally was also higher using the reported deaths method than using the reported cases method, for all regions except SEARO (Figure 5A). The averaged percentage of the population infected was lowest in WPRO at 0.66% (95% CI: 0.59-0.75%), followed by AFRO at 3.25% (95% CI: 2.75-3.84%), SEARO at 8.11% (95% CI: 7.05-9.24%), EMRO at 13.67% (95% CI: 11.83-15.75%), EURO at 25.48% (95% CI: 22.76-28.40%), and highest in AMRO at 41.91% (95% CI: 37.95-46.09%) (Figure 5B). Globally, the averaged percentage of the population infected was estimated at 12.56% (95% CI: 11.17-14.05%).

**Figure 5:**
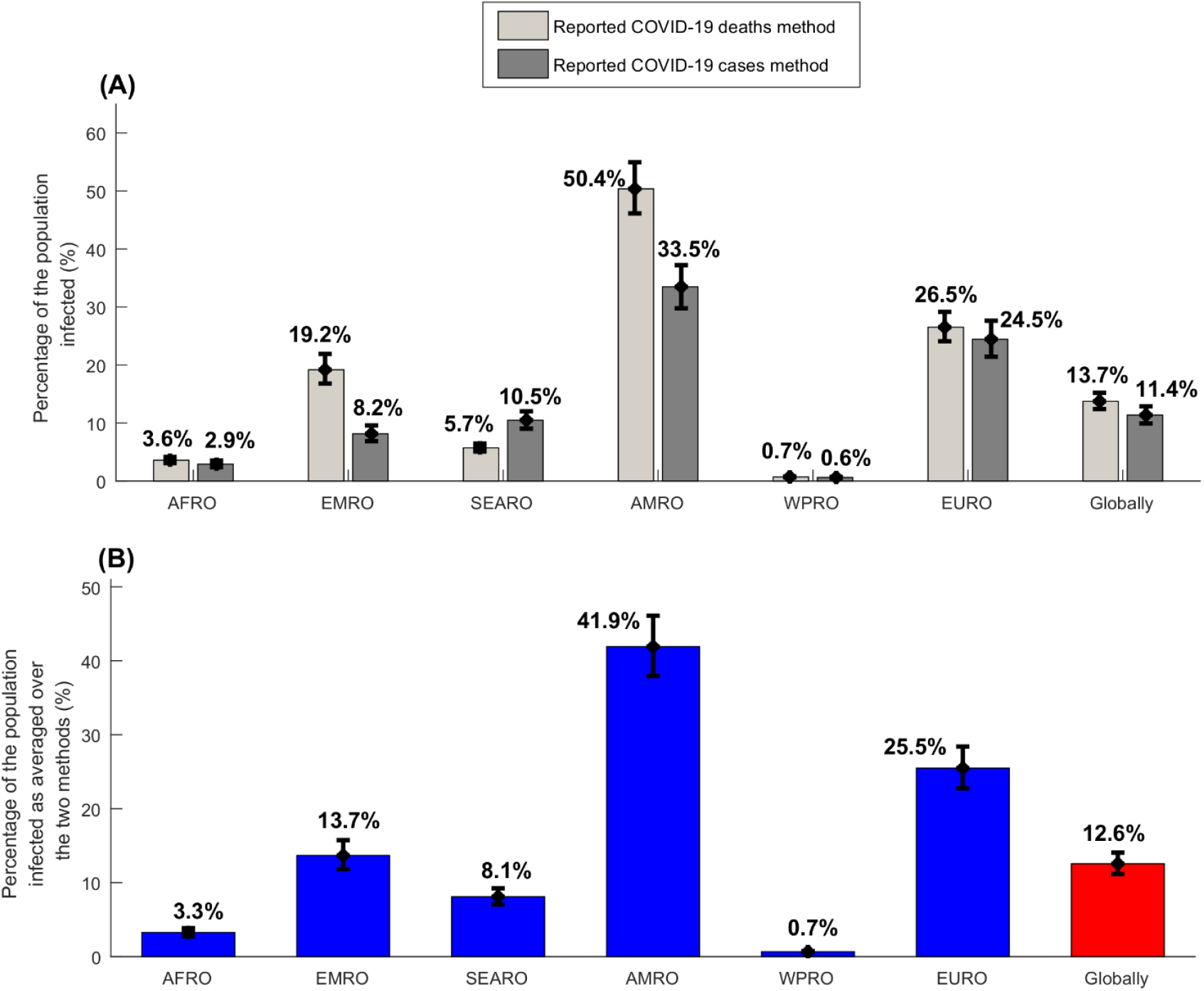
Estimated percentages of populations infected across WHO regions and globally. A) Estimated percentages of populations infected using the reported COVID-19 deaths method and the reported COVID-19 cases method. B) Estimated percentages of the population infected as averaged over the two methods. WHO regions include the African Region (AFRO), the Eastern Mediterranean Region (EMRO), the South-East Asia Region (SEARO), the Region of the Americas (AMRO), the Western Pacific Region (WPRO), and the European Region (EURO) (Figure 1).

As of December 31, 2020, 1.79 million COVID-19 deaths had been reported, but the percentage of the global population infected was estimated at only 12.56% (95% CI: 11.17-14.05%). If the global population is to reach herd immunity with no vaccination, conservatively estimated at 60-70% infection exposure [10, 35, 36], then a cumulative total of 8.18 million (95% CI: 7.30-9.18 million) COVID-19 deaths would occur by the time herd immunity is reached. Also, by then, a cumulative total of 163.67 million (95% CI: 148.12-179.51) acute-care hospitalizations, 33.01 million (95% CI: 30.52-35.70) ICU hospitalizations, 50.23 million (95% CI: 46.24-54.67) severe COVID-19 cases, and 17.62 million (95% CI: 16.36-18.97) COVID-19 critical cases would have occurred.

## DISCUSSION

The above results suggest that only 13% of the world’s population has been infected by SARS-CoV-2, even though an entire year has already passed since the epidemic emerged in Wuhan, Hubei Province, China, in December, 2019 [37, 38]. This demonstrates that the overall global population remains far below the herd immunity threshold, estimated at 60-70% infection exposure (if not more for the mutant viral strains) [10, 35, 36], and is still at risk of repeated epidemic waves of infection, with all that entails in terms of disease burden and social and economic disruption. This finding highlights the urgent need to accelerate COVID-19 vaccination to avert global expansion of this infection.

Though overall exposure to this infection remains relatively low, there are immense variations by region and country, and the Americas appear to have already reached ∼40% exposure, over 60-fold higher than the Western Pacific Region (<1%). The average incidence rate experienced since epidemic onset varies similarly and is highest in the Americas, at 132 per 10,000 person-weeks and lowest in the Western Pacific Region at only 1 per 10,000 person-weeks. These findings demonstrate strikingly high variability in the intensity of national epidemics during the first year since this infection’s introduction. It remains to be seen whether this variability reflects different national responses to the epidemics, and/or clinical or biological cofactors that make some populations more affected than others.

Even though the same *age-stratified* infection morbidity and mortality rates were used in generating estimates for all countries, total-population morbidity and mortality rates varied hugely among countries and by region, only because of differences in population age structures. For instance, the infection fatality rate in the European Region of 20 per 10,000 infections was nearly 6-fold higher than that in the African Region at <4 per 10,000 infections. Similarly, infection hospitalization and severity rates varied enormously. These findings may explain apparent variability in the severity of this infection across countries and regions, and suggest that the disease burden could be substantially lower in countries with younger demographics, such as the African or Eastern Mediterranean Regions, as suggested earlier [23].

Notwithstanding this global variability, the above results corroborate the vast disease burden that this infection can cause. Nearly two million deaths have been confirmed worldwide as of December 31, 2020 [27], though only 13% of the global population has been infected. If we were to adopt today a herd immunity approach to dealing with this pandemic worldwide, that is, achieving the herd immunity threshold without vaccination, the pandemic would cause a total of 8 million COVID-19 deaths, 68 million COVID-19 severe and critical disease cases, and 197 million hospitalizations. These findings affirm the wisdom of epidemic suppression approaches adopted in most countries to tackle their respective epidemics [39].

Despite the high potential disease burden, the above-estimated infection morbidity and mortality rates are still substantially lower than those estimated earlier in the epidemic [6, 40-44]. Globally, out of every 10,000 infections, only 11 would result in COVID-19 deaths. Out of every 1,000 infections, only 6 would be severe and only 2 would be critical per WHO classification [13]. Nineteen would be hospitalized in an acute-care bed and 4 in an ICU bed.

This study has limitations. In essence, it was based on age-stratified infection morbidity and mortality rates and the infection detection rate, estimated for a well-characterized and thoroughly investigated national epidemic, in which about half the population has already been infected [8-12, 15-21]. While the epidemic of Qatar is perhaps the world’s most advanced SARS-CoV-2 epidemic [8-12, 15-21], the extent to which fundamental infection metrics estimated with precision for one country, even if they are primarily determined by the basic biology of this infection, can be extrapolated to other countries, remains unknown. It is reasonable that these metrics could be affected by myriad factors, such as clinical or biological variations in human populations and circulating viral strains, the nature of COVID-19 responses, coverage of SARS-CoV-2 testing, and quality of reporting of cases and deaths. For instance, infection exposure is likely overestimated for countries with higher testing coverage and underestimated for countries with lower coverage. However, to make these estimates as realistic as possible, we used two independent methods with different input data to estimate infection exposure, to minimize the effect of any potential bias in either method or its data input. We also adjusted estimates for variation in healthcare access and quality by utilizing the Global Burden of Disease study’s Healthcare Access and Quality Index for each country [28]. Still, the provided estimates should be seen as provisional, crude estimates for the purpose of providing a broad understanding of the global epidemiology of this infection and to guide the global COVID-19 response.

## CONCLUSION

Only 13% of the global population has been infected with SARS-CoV-2 suggesting that the world’s population remains far below the herd immunity threshold and at risk of repeated epidemic waves of infection. Nevertheless, global epidemiology demonstrates immense regional variation in both infection exposure and SARS-CoV-2 morbidity and mortality rates. While the pandemic’s expansion in nations with young populations could lead to a relatively milder disease burden than current expectations, this infection has the potential to cause nearly ten million COVID-19 deaths and 200 million hospitalizations worldwide, if its transmission is left unchecked and vaccination scale-up continues to lag far behind global needs.

## Data Availability

All data generated or analyzed during this study are included in this article and its Supplementary Information file.

## Acknowledgements

The authors are also grateful for support provided by the Biomedical Research Program and the Biostatistics, Epidemiology, and Biomathematics Research Core at Weill Cornell Medicine-Qatar. Developed mathematical models were made possible by NPRP grant number 9-040-3-008 and NPRP grant number 12S-0216-190094 from the Qatar National Research Fund (a member of Qatar Foundation; https://www.qnrf.org). GM acknowledges support by UK Research and Innovation as part of the Global Challenges Research Fund, grant number ES/P010873/1. Statements made herein are solely the responsibility of the authors. Funders had no role in study design, data collection and analysis, decision to publish, or preparation of the manuscript.

## Author contributions

HHA constructed and parameterized the mathematical model, conducted the mathematical modeling analyses, and co-wrote the first draft of the manuscript. HC and GM contributed to the parameterization of the model. LJA conceived and led the design of the study and model, conduct of analyses, and co-wrote the first draft of the manuscript. All authors contributed to discussions and interpretation of the results and to the writing of the manuscript. All authors have read and approved the final manuscript.

## Supporting Information

**Table S1:** Estimates for key SARS-CoV-2 epidemiologic indicators for countries and territories with a population of at least one million [1] in the WHO African Region (AFRO). Classification of infection severity and criticality was per WHO infection severity classifications [2]. Classification of COVID-19 death was also per WHO guidelines [3].

| African Region | Total population [1]<sup>a</sup> | HAQ Index [4]<sup>b</sup> | Infection acute-care bed hospitalization rate (per 1,000 infections) | Infection ICU bed hospitalization rate (per 1,000 infections) | Infection severity rate (per 1,000 infections) | Infection criticality rate (per 1,000 infections) | Infection fatality rate (per 10,000 infections) | Infection detection rate (%) | Incidence rate since epidemic onset (per 10,000 person-weeks)<sup>c</sup> | Proportion of the population infected (%)<sup>d</sup> |
| --- | --- | --- | --- | --- | --- | --- | --- | --- | --- | --- |
| Country | Thousands | N (95% CI) | N (95% CI) | N (95% CI) | N (95% CI) | N (95% CI) | N (95% CI) | N (95% CI) | N (95% CI) | N (95% CI) |
| Algeria | 43,851,043 | 63.1 (59.4-66.4) | 16.7 (16.3-16.9) | 3.2 (3.2-3.2) | 4.9 (4.8-5.0) | 1.7 (1.7-1.7) | 7.8 (7.4-8.2) | 8.5 (7.4-9.9) | 9.6 (8.4-10.9) | 4.1 (3.6-4.7) |
| Angola | 32,866,268 | 33.4 (25.5-40.4) | 11.6 (11.3-11.8) | 1.5 (1.5-1.5) | 2.2 (2.2-2.2) | 0.7 (0.7-0.7) | 2.6 (2.5-2.8) | 4.5 (3.2-6.0) | 3.6 (2.6-4.8) | 1.4 (1.1-2.0) |
| Benin | 12,123,198 | 30.8 (27.8-34.0) | 12.7 (12.4-12.9) | 1.9 (1.9-1.9) | 2.8 (2.8-2.9) | 0.9 (0.9-0.9) | 3.8 (3.6-4.0) | 4.2 (3.5-5.1) | 1.2 (1.0-1.4) | 0.5 (0.4-0.6) |
| Botswana | 2,351,625 | 51.5 (40.8-69.2) | 14.4 (14.1-14.6) | 2.4 (2.4-2.4) | 3.7 (3.6-3.8) | 1.2 (1.2-1.2) | 5.0 (4.8-5.3) | 7.0 (5.1-10.3) | 13.9 (9.5-19.6) | 5.3 (3.6-7.3) |
| Burkina Faso | 20,903,278 | 30.1 (27.0-33.3) | 11.8 (11.5-12.0) | 1.6 (1.6-1.6) | 2.3 (2.3-2.4) | 0.7 (0.7-0.7) | 2.8 (2.6-2.9) | 4.1 (3.4-5.0) | 1.5 (1.2-1.8) | 0.6 (0.5-0.7) |
| Burundi | 11,890,781 | 27.4 (23.1-32.1) | 11.7 (11.4-11.8) | 1.5 (1.5-1.5) | 2.2 (2.2-2.3) | 0.7 (0.7-0.7) | 2.7 (2.5-2.8) | 3.7 (2.9-4.8) | 0.3 (0.2-0.3) | 0.1 (0.1-0.1) |
| Cameroon | 26,545,864 | 31.9 (26.9-37.5) | 12.2 (11.9-12.3) | 1.7 (1.7-1.7) | 2.5 (2.5-2.6) | 0.8 (0.8-0.8) | 3.1 (2.9-3.2) | 4.3 (3.4-5.6) | 5.0 (3.9-6.4) | 2.1 (1.7-2.7) |
| Cent. Afr. Rep. | 4,829,764 | 18.6 (13.1-24.4) | 12.1 (11.8-12.3) | 1.7 (1.7-1.7) | 2.5 (2.4-2.5) | 0.8 (0.8-0.8) | 3.2 (3.0-3.4) | 2.5 (1.6-3.6) | 6.0 (4.1-9.2) | 2.5 (1.7-3.7) |
| Chad | 16,425,859 | 25.4 (21.9-29.0) | 11.6 (11.3-11.8) | 1.5 (1.5-1.6) | 2.2 (2.2-2.3) | 0.7 (0.7-0.7) | 2.8 (2.7-2.9) | 3.4 (2.7-4.3) | 1.2 (1.0-1.5) | 0.5 (0.4-0.6) |
| Congo | 5,518,092 | 34.1 (28.4-40.4) | 12.6 (12.3-12.8) | 1.8 (1.8-1.8) | 2.7 (2.7-2.8) | 0.8 (0.8-0.8) | 3.2 (3.0-3.4) | 4.6 (3.5-6.0) | 5.6 (4.3-7.2) | 2.3 (1.8-2.9) |
| Côte d'Ivoire | 26,378,275 | 27.3 (24.2-31.1) | 12.4 (12.1-12.6) | 1.7 (1.7-1.8) | 2.6 (2.6-2.7) | 0.8 (0.8-0.8) | 3.2 (3.0-3.4) | 3.7 (3.0-4.6) | 3.3 (2.7-4.1) | 1.4 (1.1-1.7) |
| D. Rep. Congo | 89,561,404 | 29.6 (25.7-33.7) | 12.3 (12.0-12.5) | 1.8 (1.7-1.8) | 2.6 (2.5-2.6) | 0.8 (0.8-0.8) | 3.4 (3.3-3.6) | 4.0 (3.2-5.0) | 1.3 (1.1-1.6) | 0.6 (0.4-0.7) |
| Equ. Guinea | 1,402,985 | 49.3 (38.3-62.0) | 11.9 (11.6-12.0) | 1.6 (1.5-1.6) | 2.4 (2.4-2.4) | 0.7 (0.7-0.7) | 2.7 (2.5-2.8) | 6.7 (4.8-9.2) | 22.9 (16.4-31.8) | 9.0 (6.6-12.3) |
| Eritrea | 3,546,427 | 27.6 (23.7-31.3) | 13.6 (13.2-13.8) | 2.2 (2.2-2.3) | 3.3 (3.2-3.3) | 1.1 (1.1-1.1) | 5.0 (4.8-5.3) | 3.7 (3.0-4.7) | 1.2 (0.9-1.5) | 0.5 (0.4-0.6) |
| Ethiopia | 114,963,583 | 28.1 (24.3-32.2) | 12.8 (12.5-13.0) | 2.0 (1.9-2.0) | 2.9 (2.8-2.9) | 0.9 (0.9-0.9) | 4.1 (3.8-4.3) | 3.8 (3.0-4.8) | 5.0 (4.0-6.2) | 2.1 (1.6-2.6) |
| Eswatini | 1,160,164 | 40.5 (30.4-52.2) | 13.1 (12.8-13.3) | 2.1 (2.1-2.1) | 3.1 (3.0-3.1) | 1.0 (1.0-1.0) | 4.6 (4.3-4.8) | 5.5 (3.8-7.8) | 39.1 (26.8-57.1) | 15.0 (10.6-21.2) |
| Gabon | 2,225,728 | 40.4 (35.0-46.1) | 13.3 (13.0-13.5) | 2.1 (2.0-2.1) | 3.1 (3.1-3.2) | 1.0 (1.0-1.0) | 4.1 (3.9-4.3) | 5.5 (4.4-6.9) | 13.6 (10.8-17.1) | 5.5 (4.4-6.8) |
| Gambia | 2,416,664 | 35.7 (32.3-39.3) | 11.9 (11.6-12.1) | 1.6 (1.6-1.6) | 2.4 (2.3-2.4) | 0.7 (0.7-0.7) | 2.9 (2.7-3.1) | 4.8 (4.0-5.9) | 12.9 (10.7-15.4) | 5.1 (4.3-6.1) |
| Ghana | 31,072,945 | 39.3 (36.0-43.4) | 13.3 (13.0-13.5) | 2.0 (2.0-2.0) | 3.1 (3.0-3.1) | 0.9 (0.9-0.9) | 3.7 (3.5-3.9) | 5.3 (4.5-6.5) | 5.6 (4.6-6.6) | 2.3 (1.9-2.7) |
| Guinea | 13,132,792 | 26.4 (22.6-30.2) | 12.2 (11.9-12.4) | 1.7 (1.7-1.7) | 2.5 (2.5-2.6) | 0.8 (0.8-0.8) | 3.2 (3.1-3.4) | 3.6 (2.8-4.5) | 4.2 (3.3-5.3) | 1.7 (1.4-2.2) |
| Guinea-Bissau | 1,967,998 | 23.4 (20.2-26.8) | 12.3 (12.0-12.5) | 1.7 (1.7-1.7) | 2.6 (2.5-2.6) | 0.8 (0.8-0.8) | 3.1 (2.9-3.3) | 3.2 (2.5-4.0) | 7.4 (5.9-9.3) | 2.9 (2.3-3.7) |
| Kenya | 53,771,300 | 39.5 (35.0-43.9) | 12.2 (11.9-12.4) | 1.6 (1.6-1.7) | 2.5 (2.5-2.6) | 0.7 (0.7-0.7) | 2.9 (2.7-3.0) | 5.3 (4.4-6.5) | 9.9 (8.1-12.0) | 4.0 (3.3-4.9) |
| Lesotho | 2,142,252 | 32.0 (24.6-40.3) | 14.7 (14.3-14.9) | 2.5 (2.5-2.6) | 3.9 (3.8-3.9) | 1.3 (1.3-1.3) | 5.7 (5.4-6.0) | 4.3 (3.1-6.0) | 6.6 (4.7-9.2) | 2.1 (1.5-3.0) |
| Liberia | 5,057,677 | 32.2 (29.3-35.4) | 12.9 (12.6-13.1) | 1.9 (1.9-2.0) | 2.9 (2.9-2.9) | 0.9 (0.9-0.9) | 3.8 (3.6-4.0) | 4.4 (3.7-5.3) | 2.9 (2.4-3.4) | 1.2 (1.0-1.4) |
| Madagascar | 27,691,019 | 29.6 (24.3-35.1) | 12.7 (12.4-12.9) | 1.8 (1.8-1.9) | 2.8 (2.7-2.8) | 0.9 (0.9-0.9) | 3.5 (3.4-3.7) | 4.0 (3.0-5.2) | 3.1 (2.3-4.0) | 1.2 (0.9-1.6) |
| Malawi | 19,129,955 | 32.2 (26.9-38.1) | 11.9 (11.6-12.1) | 1.6 (1.6-1.7) | 2.4 (2.4-2.4) | 0.7 (0.7-0.7) | 3.0 (2.9-3.2) | 4.4 (3.4-5.7) | 2.5 (1.9-3.2) | 1.0 (0.8-1.2) |
| Mali | 20,250,834 | 34.9 (29.9-40.1) | 11.7 (11.4-11.9) | 1.5 (1.5-1.6) | 2.2 (2.2-2.3) | 0.7 (0.7-0.7) | 2.7 (2.6-2.9) | 4.7 (3.7-6.0) | 3.3 (2.6-4.1) | 1.3 (1.0-1.6) |
| Mauritania | 4,649,660 | 40.6 (35.0-47.5) | 12.9 (12.6-13.1) | 1.9 (1.9-1.9) | 2.9 (2.8-2.9) | 0.9 (0.9-0.9) | 3.7 (3.5-3.9) | 5.5 (4.4-7.1) | 17.2 (13.6-21.9) | 6.9 (5.5-8.7) |
| Mauritius | 1,271,767 | 68.7 (65.5-71.9) | 23.0 (22.4-23.4) | 5.3 (5.3-5.4) | 8.2 (8.1-8.3) | 3.0 (2.9-3.0) | 13.9 (13.3-14.7) | 9.3 (8.2-10.7) | 1.1 (0.9-1.2) | 0.4 (0.4-0.5) |
| Mozambique | 31,255,435 | 30.0 (25.3-35.0) | 12.1 (11.8-12.3) | 1.7 (1.7-1.7) | 2.5 (2.4-2.5) | 0.8 (0.8-0.8) | 3.2 (3.1-3.4) | 4.1 (3.2-5.2) | 2.5 (1.9-3.2) | 1.0 (0.8-1.3) |
| Namibia | 2,540,916 | 44.6 (36.4-56.2) | 13.3 (12.9-13.5) | 2.1 (2.0-2.1) | 3.1 (3.1-3.2) | 1.0 (1.0-1.0) | 4.2 (4.0-4.4) | 6.0 (4.5-8.4) | 31.1 (22.5-42.7) | 12.1 (8.9-16.2) |
| Niger | 24,206,636 | 28.4 (23.9-33.1) | 11.8 (11.5-11.9) | 1.6 (1.6-1.6) | 2.3 (2.2-2.3) | 0.7 (0.7-0.7) | 2.9 (2.7-3.0) | 3.8 (3.0-4.9) | 0.9 (0.7-1.2) | 0.4 (0.3-0.5) |
| Nigeria | 206,139,587 | 41.9 (37.2-47.3) | 12.4 (12.1-12.6) | 1.7 (1.7-1.7) | 2.6 (2.5-2.6) | 0.8 (0.8-0.8) | 3.0 (2.9-3.2) | 5.7 (4.6-7.1) | 1.9 (1.6-2.3) | 0.8 (0.7-1.0) |
| Rwanda | 12,952,209 | 36.0 (31.6-40.5) | 12.8 (12.5-13.0) | 1.9 (1.8-1.9) | 2.8 (2.8-2.9) | 0.9 (0.9-0.9) | 3.5 (3.3-3.7) | 4.9 (3.9-6.0) | 2.5 (2.0-3.1) | 1.0 (0.8-1.3) |
| Senegal | 16,743,930 | 31.1 (28.3-33.8) | 12.5 (12.2-12.7) | 1.8 (1.8-1.8) | 2.7 (2.6-2.7) | 0.8 (0.8-0.8) | 3.5 (3.3-3.7) | 4.2 (3.5-5.0) | 5.9 (5.0-7.0) | 2.5 (2.1-3.0) |
| Sierra Leone | 7,976,985 | 31.0 (27.4-34.5) | 12.5 (12.2-12.7) | 1.8 (1.8-1.8) | 2.7 (2.6-2.7) | 0.8 (0.8-0.8) | 3.4 (3.2-3.5) | 4.2 (3.4-5.1) | 2.2 (1.8-2.7) | 0.9 (0.7-1.1) |
| South Africa | 59,308,690 | 49.7 (47.2-52.4) | 15.8 (15.4-16.1) | 2.8 (2.8-2.8) | 4.4 (4.3-4.5) | 1.4 (1.4-1.4) | 6.2 (5.9-6.5) | 6.7 (5.9-7.8) | 97.3 (83.1-114.3) | 33.7 (29.7-38.2) |
| South Sudan | 11,193,729 | 26.8 (21.0-33.1) | 12.8 (12.5-13.0) | 1.9 (1.9-2.0) | 2.8 (2.8-2.9) | 0.9 (0.9-0.9) | 3.9 (3.7-4.1) | 3.6 (2.6-4.9) | 1.7 (1.2-2.3) | 0.7 (0.5-0.9) |
| Togo | 8,278,737 | 32.0 (28.7-35.6) | 12.6 (12.3-12.8) | 1.8 (1.8-1.8) | 2.7 (2.7-2.8) | 0.8 (0.8-0.8) | 3.3 (3.1-3.4) | 4.3 (3.6-5.3) | 2.2 (1.8-2.7) | 0.9 (0.8-1.1) |
| Uganda | 45,741,000 | 31.4 (27.2-35.6) | 11.2 (10.9-11.4) | 1.4 (1.4-1.4) | 2.0 (2.0-2.0) | 0.6 (0.6-0.6) | 2.3 (2.2-2.4) | 4.2 (3.4-5.3) | 3.2 (2.6-4.0) | 1.3 (1.1-1.6) |
| Tanzania | 59,734,213 | 33.9 (30.0-38.4) | 12.1 (11.8-12.3) | 1.7 (1.6-1.7) | 2.5 (2.4-2.5) | 0.8 (0.7-0.8) | 3.0 (2.9-3.2) | 4.6 (3.7-5.7) | 0.1 (0.1-0.1) | 0.0 (0.0-0.0) |
| Zambia | 18,383,956 | 29.0 (23.0-35.4) | 11.4 (11.1-11.6) | 1.5 (1.4-1.5) | 2.1 (2.1-2.2) | 0.6 (0.6-0.6) | 2.5 (2.4-2.6) | 3.9 (2.9-5.3) | 6.8 (5.1-9.2) | 2.8 (2.1-3.7) |
| Zimbabwe | 14,862,927 | 31.2 (25.8-37.0) | 12.3 (12.0-12.5) | 1.8 (1.7-1.8) | 2.6 (2.6-2.7) | 0.8 (0.8-0.8) | 3.4 (3.3-3.6) | 4.2 (3.2-5.5) | 5.7 (4.4-7.4) | 2.3 (1.8-3.0) |
| **AFRO** | **1,118,418,151** | **35.6 (31.3-40.2)** | **12.7 (12.3-12.8)** | **1.8 (1.8-1.9)** | **2.8 (2.7-2.8)** | **0.9 (0.9-0.9)** | **3.6 (3.4-3.8)** | **4.8 (3.9-6.0)** | **8.7 (7.2-10.5)** | **3.3 (2.8-3.8)** |

Cent Afr Rep, Central African Republic. CI, credible interval. D Rep Congo, Democratic Republic of the Congo. Equ Guinea, Equatorial Guinea. HAQ, Healthcare Access and Quality. N, number. Tanzania, United Republic of Tanzania.
<sup>a</sup>The population size and the demographic age-structure were extracted from the United Nations World Population Prospects database [1].
<sup>b</sup>HAQ Index was extracted from the Global Burden of Disease study [4].
<sup>c</sup>The estimate for the incidence rate was based on the average of the two estimates generated by the reported COVID-19 deaths method and the reported COVID-19 cases method.
<sup>d</sup>The estimate for the proportion of the population infected was based on the average of the two estimates generated by the reported COVID-19 deaths method and the reported COVID-19 cases method.

**Table S2:** Estimates for key SARS-CoV-2 epidemiologic indicators for countries and territories with a population of at least one million [1] in the WHO Region of the Americas (AMRO). Classification of infection severity and criticality was per WHO infection severity classifications [2]. Classification of COVID-19 death was also per WHO guidelines [3].

| Region of the Americas | Total population [1]<sup>a</sup> | HAQ Index [4]<sup>b</sup> | Infection acute-care bed hospitalization rate (per 1,000 infections) | Infection ICU bed hospitalization rate (per 1,000 infections) | Infection severity rate (per 1,000 infections) | Infection criticality rate (per 1,000 infections) | Infection fatality rate (per 10,000 infections) | Infection detection rate (%) | Incidence rate since epidemic onset (per 10,000 person-weeks)<sup>c</sup> | Proportion of the population infected (%)<sup>d</sup> |
| --- | --- | --- | --- | --- | --- | --- | --- | --- | --- | --- |
| Country | Thousands | N (95% CI) | N (95% CI) | N (95% CI) | N (95% CI) | N (95% CI) | N (95% CI) | N (95% CI) | N (95% CI) | N (95% CI) |
| Argentina | 45,195,777 | 68.1 (65.8-70.1) | 20.4 (19.9-20.7) | 4.8 (4.7-4.8) | 7.0 (6.9-7.1) | 2.7 (2.7-2.7) | 13.4 (12.7-14.0) | 9.2 (8.2-10.5) | 142.5 (123.1-165.1) | 45.6 (40.9-50.5) |
| Bolivia | 11,673,029 | 48.8 (43.5-54.0) | 16.6 (16.2-16.8) | 3.4 (3.4-3.4) | 5.0 (4.9-5.1) | 1.8 (1.8-1.8) | 8.9 (8.5-9.4) | 6.6 (5.4-8.1) | 101.6 (80.6-128.5) | 33.5 (27.9-39.9) |
| Brazil | 212,559,409 | 63.8 (62.3-64.9) | 19.8 (19.3-20.1) | 4.3 (4.3-4.4) | 6.6 (6.5-6.7) | 2.4 (2.3-2.4) | 11.2 (10.6-11.8) | 8.6 (7.8-9.7) | 152.9 (133.4-175.2) | 48.6 (44.0-53.3) |
| Canada | 37,742,157 | 93.8 (92.8-94.8) | 26.8 (26.1-27.2) | 7.2 (7.1-7.3) | 10.5 (10.4-10.7) | 4.2 (4.1-4.2) | 21.3 (20.3-22.4) | 12.7 (11.6-14.1) | 35.4 (32.2-38.7) | 15.7 (14.4-17.1) |
| Chile | 19,116,209 | 77.9 (72.3-83.7) | 21.9 (21.4-22.3) | 5.2 (5.1-5.2) | 7.8 (7.7-7.9) | 2.9 (2.9-2.9) | 14.3 (13.6-15.1) | 10.5 (9.0-12.5) | 123.5 (101.8-151.0) | 40.5 (34.9-46.7) |
| Colombia | 50,882,884 | 68.5 (65.8-70.9) | 19.1 (18.6-19.3) | 4.1 (4.0-4.1) | 6.2 (6.1-6.3) | 2.2 (2.2-2.2) | 10.5 (10.0-11.1) | 9.3 (8.2-10.6) | 152.5 (130.2-179.4) | 46.5 (41.6-51.8) |
| Costa Rica | 5,094,114 | 73.7 (71.2-76.0) | 20.4 (19.9-20.8) | 4.6 (4.5-4.6) | 7.0 (6.8-7.1) | 2.5 (2.5-2.6) | 12.2 (11.6-12.8) | 10.0 (8.9-11.3) | 85.5 (74.6-97.6) | 30.5 (27.2-33.9) |
| Cuba | 11,326,616 | 75.5 (73.5-77.7) | 25.4 (24.8-25.8) | 6.6 (6.5-6.6) | 9.7 (9.5-9.8) | 3.7 (3.7-3.8) | 18.8 (17.9-19.8) | 10.2 (9.2-11.6) | 1.9 (1.7-2.1) | 0.8 (0.7-0.9) |
| Dom. Rep. | 10,847,904 | 61.2 (57.3-65.6) | 17.3 (16.9-17.6) | 3.5 (3.5-3.6) | 5.3 (5.2-5.4) | 1.9 (1.9-1.9) | 8.9 (8.5-9.4) | 8.3 (7.1-9.8) | 45.2 (38.4-53.0) | 17.8 (15.4-20.6) |
| Ecuador | 17,643,060 | 62.2 (59.5-64.6) | 17.3 (16.9-17.6) | 3.5 (3.5-3.5) | 5.3 (5.2-5.4) | 1.9 (1.9-1.9) | 8.9 (8.4-9.3) | 8.4 (7.4-9.6) | 122.9 (104.5-145.7) | 37.2 (33.3-41.4) |
| El Salvador | 6,486,201 | 63.2 (58.9-67.2) | 17.9 (17.4-18.1) | 3.8 (3.8-3.9) | 5.7 (5.6-5.8) | 2.1 (2.1-2.1) | 10.2 (9.7-10.7) | 8.6 (7.3-10.0) | 28.9 (24.9-33.5) | 11.1 (9.7-12.8) |
| Guatemala | 17,915,567 | 51.5 (45.3-57.7) | 14.2 (13.8-14.4) | 2.5 (2.5-2.5) | 3.7 (3.6-3.7) | 1.3 (1.3-1.3) | 5.9 (5.6-6.2) | 7.0 (5.6-8.6) | 49.7 (39.9-61.7) | 18.3 (15.1-22.2) |
| Haiti | 11,402,533 | 32.1 (26.6-37.8) | 14.8 (14.4-15.0) | 2.6 (2.6-2.6) | 3.9 (3.9-4.0) | 1.3 (1.3-1.3) | 5.9 (5.6-6.2) | 4.3 (3.3-5.6) | 4.0 (3.1-5.2) | 1.6 (1.3-2.1) |
| Honduras | 9,904,608 | 46.5 (40.1-53.1) | 14.5 (14.2-14.7) | 2.5 (2.5-2.6) | 3.8 (3.8-3.9) | 1.3 (1.3-1.3) | 5.9 (5.6-6.2) | 6.3 (5.0-7.9) | 63.4 (49.5-81.2) | 23.3 (18.7-28.8) |
| Jamaica | 2,961,161 | 62.0 (56.8-67.3) | 19.1 (18.6-19.4) | 4.1 (4.1-4.2) | 6.2 (6.1-6.3) | 2.3 (2.2-2.3) | 10.9 (10.4-11.5) | 8.4 (7.1-10.0) | 14.1 (11.9-16.6) | 5.8 (4.9-6.8) |
| Mexico | 128,932,753 | 66.3 (64.9-67.4) | 17.7 (17.2-17.9) | 3.6 (3.5-3.6) | 5.5 (5.4-5.5) | 1.9 (1.9-1.9) | 9.0 (8.5-9.4) | 9.0 (8.1-10.1) | 176.5 (149.9-212.4) | 44.0 (40.4-47.8) |
| Nicaragua | 6,624,554 | 61.2 (57.0-65.4) | 15.5 (15.2-15.8) | 2.8 (2.8-2.9) | 4.3 (4.3-4.4) | 1.5 (1.5-1.5) | 6.6 (6.3-7.0) | 8.3 (7.1-9.8) | 4.2 (3.7-4.9) | 1.7 (1.5-2.0) |
| Panama | 4,314,768 | 68.3 (64.6-71.9) | 18.3 (17.8-18.6) | 3.9 (3.9-3.9) | 5.8 (5.8-5.9) | 2.1 (2.1-2.1) | 10.3 (9.8-10.8) | 9.2 (8.0-10.7) | 233.1 (188.9-292.8) | 62.6 (54.9-70.9) |
| Paraguay | 7,132,530 | 56.7 (53.1-60.2) | 16.3 (15.9-16.6) | 3.2 (3.1-3.2) | 4.8 (4.7-4.9) | 1.7 (1.7-1.7) | 7.8 (7.4-8.2) | 7.7 (6.6-9.0) | 59.0 (50.3-69.0) | 22.2 (19.2-25.4) |
| Peru | 32,971,846 | 64.3 (59.2-69.4) | 18.6 (18.1-18.9) | 3.9 (3.9-4.0) | 6.0 (5.9-6.1) | 2.1 (2.1-2.2) | 10.2 (9.7-10.7) | 8.7 (7.4-10.4) | 223.0 (170.9-311.6) | 56.1 (48.3-64.8) |
| Puerto Rico | 2,860,840 | 82.7 (80.2-85.0) | 28.8 (28.1-29.3) | 8.2 (8.1-8.3) | 11.7 (11.6-11.9) | 4.8 (4.7-4.8) | 25.1 (23.9-26.4) | 11.2 (10.0-12.7) | 57.5 (50.7-64.8) | 21.3 (19.1-23.6) |
| Trin. and Tob. | 1,399,491 | 64.3 (60.7-67.5) | 22.0 (21.5-22.4) | 5.0 (5.0-5.0) | 7.7 (7.6-7.8) | 2.8 (2.7-2.8) | 13.0 (12.4-13.7) | 8.7 (7.6-10.1) | 13.2 (11.5-15.1) | 5.4 (4.7-6.1) |
| USA | 331,002,647 | 88.7 (88.0-89.4) | 25.3 (24.7-25.7) | 6.6 (6.6-6.7) | 9.7 (9.5-9.9) | 3.8 (3.8-3.8) | 19.4 (18.5-20.4) | 12.0 (11.0-13.3) | 137.7 (121.9-155.2) | 49.4 (45.2-53.6) |
| Uruguay | 3,473,727 | 71.0 (68.9-73.0) | 23.2 (22.6-23.6) | 6.1 (6.0-6.2) | 8.7 (8.6-8.9) | 3.5 (3.5-3.5) | 18.5 (17.6-19.4) | 9.6 (8.6-10.9) | 9.2 (8.1-10.2) | 3.7 (3.3-4.1) |
| Venezuela | 28,435,943 | 67.8 (63.6-71.8) | 18.4 (17.9-18.6) | 3.8 (3.7-3.8) | 5.8 (5.7-5.8) | 2.0 (2.0-2.0) | 9.3 (8.8-9.7) | 9.2 (7.9-10.7) | 8.9 (7.6-10.2) | 3.6 (3.1-4.2) |
| **AMRO** | **1,017,900,328** | **73.4 (71.5-75.2)** | **21.3 (20.7-21.6)** | **5.0 (4.9-5.0)** | **7.4 (7.3-7.6)** | **2.8 (2.8-2.8)** | **13.8 (13.1-14.5)** | **9.9 (8.9-11.2)** | **131.7 (113.6-154.2)** | **41.9 (37.9-46.1)** |

CI, credible interval. Dom Rep, Dominican Republic. HAQ, Healthcare Access and Quality. N, number. Trin. and Tob., Trinidad and Tobago.
<sup>a</sup>The population size and the demographic age-structure were extracted from the United Nations World Population Prospects database [1].
<sup>b</sup>HAQ Index was extracted from the Global Burden of Disease study [4].
<sup>c</sup>The estimate for the incidence rate was based on the average of the two estimates generated by the reported COVID-19 deaths method and the reported COVID-19 cases method.
<sup>d</sup>The estimate for the proportion of the population infected was based on the average of the two estimates generated by the reported COVID-19 deaths method and the reported COVID-19 cases method.

**Table S3:**
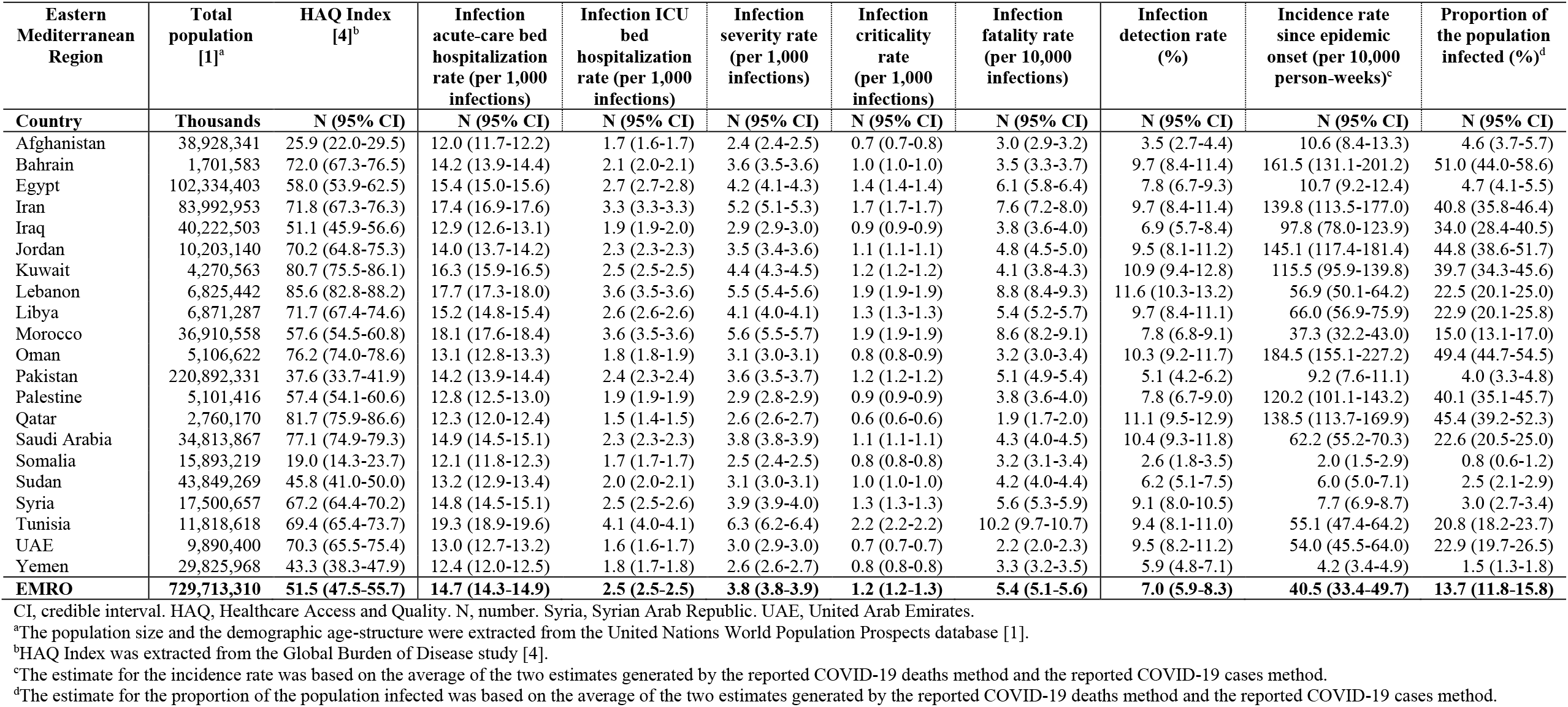
Estimates for key SARS-CoV-2 epidemiologic indicators for countries and territories with a population of at least one million [1] in the WHO Eastern Mediterranean Region (EMRO). Classification of infection severity and criticality was per WHO infection severity classifications [2]. Classification of COVID-19 death was also per WHO guidelines [3].

**Table S4:** Estimates for key SARS-CoV-2 epidemiologic indicators for countries and territories with a population of at least one million [1] in the WHO European Region (EURO). Classification of infection severity and criticality was per WHO infection severity classifications [2]. Classification of COVID-19 death was also per WHO guidelines [3].

| European Region | Total population [1]<sup>a</sup> | HAQ Index [4]<sup>b</sup> | Infection acute-care bed hospitalization rate (per 1,000 infections) | Infection ICU bed hospitalization rate (per 1,000 infections) | Infection severity rate (per 1,000 infections) | Infection criticality rate (per 1,000 infections) | Infection fatality rate (per 10,000 infections) | Infection detection rate (%) | Incidence rate since epidemic onset (per 10,000 person-weeks)<sup>c</sup> | Proportion of the population infected (%)<sup>d</sup> |
| --- | --- | --- | --- | --- | --- | --- | --- | --- | --- | --- |
| Country | Thousands | N (95% CI) | N (95% CI) | N (95% CI) | N (95% CI) | N (95% CI) | N (95% CI) | N (95% CI) | N (95% CI) | N (95% CI) |
| Albania | 2,877,800 | 63.6 (75.5-78.2) | 24.5 (23.9-24.9) | 6.1 (6.0-6.2) | 9.1 (9.0-9.3) | 3.5 (3.4-3.5) | 17.1 (16.2-17.9) | 8.6 (9.4-11.7) | 52.9 (46.6-57.6) | 20.0 (17.9-21.7) |
| Armenia | 2,963,234 | 70.7 (67.8-73.5) | 22.0 (21.4-22.3) | 5.1 (5.1-5.2) | 7.8 (7.7-7.9) | 2.9 (2.9-2.9) | 14.2 (13.6-15.0) | 9.6 (8.4-11.0) | 175.4 (147.2-210.1) | 53.4 (47.3-59.8) |
| Austria | 9,006,400 | 93.9 (92.6-95.3) | 27.7 (27.0-28.1) | 7.8 (7.7-7.8) | 11.1 (11.0-11.3) | 4.5 (4.5-4.6) | 23.9 (22.7-25.1) | 12.7 (11.5-14.2) | 81.3 (72.6-90.5) | 30.1 (27.4-32.9) |
| Azerbaijan | 10,139,175 | 65.6 (61.2-69.6) | 18.2 (17.7-18.5) | 3.5 (3.5-3.5) | 5.6 (5.5-5.7) | 1.8 (1.8-1.9) | 8.1 (7.7-8.5) | 8.9 (7.6-10.4) | 61.5 (52.1-72.3) | 23.6 (20.4-27.1) |
| Belarus | 9,449,321 | 79.0 (75.3-82.8) | 25.0 (24.4-25.4) | 6.2 (6.2-6.3) | 9.4 (9.3-9.6) | 3.6 (3.5-3.6) | 17.9 (17.0-18.8) | 10.7 (9.4-12.4) | 32.9 (28.4-37.8) | 13.2 (11.5-14.9) |
| Belgium | 11,589,616 | 92.9 (90.7-95.0) | 26.9 (26.3-27.4) | 7.6 (7.5-7.6) | 10.8 (10.6-11.0) | 4.4 (4.4-4.5) | 23.5 (22.4-24.7) | 12.6 (11.3-14.2) | 193.6 (165.5-229.3) | 57.8 (52.5-63.4) |
| Bosnia & Herz. | 3,280,815 | 72.2 (67.2-76.4) | 27.3 (26.6-27.7) | 7.0 (7.0-7.1) | 10.6 (10.4-10.8) | 4.0 (4.0-4.1) | 20.1 (19.1-21.1) | 9.8 (8.4-11.4) | 126.1 (104.9-151.7) | 41.5 (36.0-47.4) |
| Bulgaria | 6,948,445 | 77.2 (73.3-80.7) | 29.5 (28.8-30.0) | 8.3 (8.2-8.3) | 12.0 (11.8-12.2) | 4.8 (4.8-4.8) | 24.7 (23.5-26.0) | 10.4 (9.1-12.0) | 92.4 (79.4-107.3) | 32.4 (28.5-36.5) |
| Croatia | 4,105,268 | 86.9 (84.5-89.4) | 28.7 (28.0-29.2) | 8.1 (8.0-8.2) | 11.7 (11.5-11.9) | 4.8 (4.7-4.8) | 25.0 (23.8-26.2) | 11.8 (10.5-13.3) | 116.1 (100.8-133.3) | 40.0 (35.8-44.3) |
| Cyprus | 1,207,361 | 90.3 (88.8-91.8) | 23.6 (23.0-24.0) | 5.9 (5.9-6.0) | 8.8 (8.6-8.9) | 3.4 (3.3-3.4) | 16.9 (16.1-17.8) | 12.2 (11.1-13.7) | 26.2 (23.4-29.0) | 10.4 (9.3-11.4) |
| Czech Republic | 10,708,982 | 89.0 (87.5-90.4) | 28.3 (27.6-28.8) | 7.7 (7.7-7.8) | 11.3 (11.1-11.5) | 4.5 (4.4-4.5) | 22.8 (21.7-23.9) | 12.0 (10.9-13.5) | 164.8 (143.0-189.6) | 50.9 (46.1-55.7) |
| Denmark | 5,792,203 | 92.1 (89.8-94.3) | 28.2 (27.5-28.7) | 8.0 (7.9-8.0) | 11.4 (11.2-11.5) | 4.6 (4.6-4.7) | 24.0 (22.9-25.3) | 12.5 (11.2-14.1) | 39.7 (35.0-44.5) | 15.8 (14.1-17.5) |
| Estonia | 1,326,539 | 85.9 (83.6-88.3) | 27.4 (26.7-27.8) | 7.7 (7.7-7.8) | 11.1 (10.9-11.3) | 4.5 (4.5-4.6) | 24.2 (23.1-25.4) | 11.6 (10.4-13.2) | 30.7 (27.1-34.5) | 12.5 (11.1-13.8) |
| Finland | 5,540,718 | 95.9 (94.6-96.9) | 29.4 (28.7-29.9) | 8.5 (8.5-8.6) | 12.1 (12.0-12.3) | 5.0 (5.0-5.0) | 26.5 (25.2-27.8) | 13.0 (11.8-14.5) | 9.6 (8.7-10.4) | 4.5 (4.1-4.9) |
| France | 65,273,512 | 91.7 (90.3-93.1) | 27.6 (26.9-28.0) | 8.0 (7.9-8.0) | 11.2 (11.1-11.4) | 4.7 (4.6-4.7) | 25.1 (24.0-26.4) | 12.4 (11.3-13.9) | 89.1 (79.5-99.6) | 35.2 (32.1-38.4) |
| Georgia | 3,989,175 | 67.1 (62.7-71.0) | 24.4 (23.8-24.8) | 6.2 (6.1-6.2) | 9.2 (9.0-9.3) | 3.5 (3.5-3.6) | 17.8 (17.0-18.7) | 9.1 (7.8-10.6) | 146.0 (116.8-187.4) | 44.4 (38.3-51.1) |
| Germany | 83,783,945 | 92.0 (90.4-93.6) | 28.9 (28.2-29.4) | 8.5 (8.4-8.6) | 12.0 (11.8-12.2) | 5.0 (5.0-5.1) | 27.2 (25.9-28.5) | 12.4 (11.3-14.0) | 35.1 (31.5-38.8) | 15.6 (14.1-17.1) |
| Greece | 10,423,056 | 90.4 (88.8-91.9) | 28.9 (28.1-29.3) | 8.7 (8.6-8.7) | 12.1 (11.9-12.3) | 5.1 (5.1-5.2) | 28.1 (26.8-29.5) | 12.2 (11.1-13.7) | 33.2 (30.0-36.5) | 13.5 (12.3-14.8) |
| Hungary | 9,660,350 | 82.1 (79.5-84.9) | 28.1 (27.4-28.6) | 7.6 (7.6-7.7) | 11.2 (11.0-11.4) | 4.4 (4.4-4.5) | 22.6 (21.5-23.7) | 11.1 (9.9-12.7) | 99.4 (86.9-113.7) | 34.6 (31.0-38.5) |
| Ireland | 4,937,796 | 94.6 (91.8-96.8) | 23.7 (23.1-24.1) | 6.0 (5.9-6.0) | 8.8 (8.7-8.9) | 3.4 (3.3-3.4) | 16.9 (16.1-17.8) | 12.8 (11.4-14.4) | 54.9 (48.9-61.3) | 21.0 (18.9-23.1) |
| Israel | 8,655,541 | 84.8 (80.7-88.4) | 20.7 (20.2-21.0) | 5.0 (4.9-5.0) | 7.2 (7.1-7.4) | 2.8 (2.8-2.8) | 14.3 (13.6-15.1) | 11.5 (10.1-13.2) | 91.9 (78.2-107.9) | 33.2 (29.2-37.6) |
| Italy | 60,461,828 | 94.9 (93.4-96.0) | 30.0 (29.2-30.5) | 9.0 (9.0-9.1) | 12.6 (12.5-12.8) | 5.4 (5.3-5.4) | 29.1 (27.8-30.6) | 12.8 (11.6-14.3) | 90.8 (81.3-101.2) | 34.9 (31.9-38.0) |
| Kazakhstan | 18,776,707 | 69.1 (64.7-73.2) | 18.3 (17.8-18.5) | 3.7 (3.6-3.7) | 5.7 (5.6-5.8) | 2.0 (2.0-2.0) | 9.1 (8.6-9.6) | 9.3 (8.1-10.9) | 30.2 (26.0-35.0) | 11.9 (10.3-13.6) |
| Kyrgyzstan | 6,524,191 | 60.6 (58.3-62.8) | 15.3 (14.9-15.5) | 2.6 (2.6-2.6) | 4.1 (4.0-4.2) | 1.3 (1.3-1.3) | 5.6 (5.3-5.8) | 8.2 (7.3-9.4) | 54.9 (48.3-62.2) | 20.1 (17.9-22.4) |
| Latvia | 1,886,202 | 80.7 (78.0-83.3) | 28.3 (27.6-28.7) | 8.0 (7.9-8.0) | 11.5 (11.3-11.6) | 4.7 (4.6-4.7) | 24.5 (23.4-25.8) | 10.9 (9.7-12.4) | 39.0 (34.3-44.0) | 15.5 (13.8-17.3) |
| Lithuania | 2,722,291 | 80.5 (78.7-82.3) | 28.1 (27.4-28.6) | 8.0 (7.9-8.1) | 11.5 (11.3-11.6) | 4.7 (4.7-4.7) | 25.1 (24.0-26.4) | 10.9 (9.8-12.3) | 97.1 (84.0-111.5) | 33.1 (29.7-36.6) |
| Netherlands | 17,134,873 | 96.1 (94.5-97.3) | 28.2 (27.5-28.7) | 7.9 (7.8-7.9) | 11.3 (11.2-11.5) | 4.6 (4.5-4.6) | 23.7 (22.6-24.9) | 13.0 (11.8-14.5) | 89.3 (79.4-99.9) | 32.3 (29.4-35.4) |
| Norway | 5,421,242 | 96.6 (94.9-97.9) | 26.0 (25.3-26.4) | 7.0 (6.9-7.1) | 10.1 (10.0-10.3) | 4.0 (4.0-4.1) | 20.8 (19.8-21.9) | 13.1 (11.8-14.6) | 12.8 (11.5-14.1) | 5.5 (5.0-6.0) |
| Poland | 37,846,605 | 82.4 (79.7-84.6) | 27.1 (26.4-27.5) | 7.2 (7.2-7.3) | 10.7 (10.5-10.8) | 4.2 (4.2-4.2) | 21.6 (20.5-22.7) | 11.1 (9.9-12.6) | 86.5 (75.9-98.2) | 31.1 (27.9-34.5) |
| Portugal | 10,196,707 | 85.7 (84.1-87.3) | 29.7 (29.0-30.2) | 8.8 (8.7-8.9) | 12.4 (12.2-12.6) | 5.2 (5.1-5.2) | 27.8 (26.5-29.2) | 11.6 (10.5-13.0) | 78.1 (69.1-87.7) | 28.5 (25.8-31.3) |
| Rep. Moldova | 4,033,963 | 67.4 (64.5-70.4) | 22.9 (22.3-23.2) | 5.2 (5.1-5.2) | 8.1 (8.0-8.2) | 2.9 (2.8-2.9) | 13.4 (12.7-14.1) | 9.1 (8.0-10.5) | 118.5 (101.3-138.7) | 39.7 (35.1-44.7) |
| Romania | 19,237,682 | 78.3 (75.9-80.7) | 27.4 (26.7-27.9) | 7.4 (7.4-7.5) | 10.9 (10.7-11.0) | 4.3 (4.3-4.3) | 22.2 (21.1-23.3) | 10.6 (9.5-12.0) | 84.3 (73.9-95.8) | 31.1 (27.8-34.5) |
| Russia | 145,934,460 | 75.1 (67.7-81.7) | 24.8 (24.2-25.2) | 6.2 (6.2-6.3) | 9.4 (9.2-9.5) | 3.6 (3.5-3.6) | 18.0 (17.1-18.9) | 10.2 (8.4-12.2) | 45.6 (37.6-55.3) | 19.6 (16.5-23.2) |
| Serbia | 8,737,370 | 77.2 (74.9-79.3) | 27.2 (26.5-27.7) | 7.2 (7.2-7.3) | 10.7 (10.5-10.8) | 4.2 (4.1-4.2) | 21.0 (20.0-22.1) | 10.4 (9.3-11.8) | 71.9 (62.4-82.2) | 25.7 (22.9-28.6) |
| Slovakia | 5,459,643 | 83.3 (80.4-86.3) | 26.1 (25.5-26.5) | 6.6 (6.6-6.7) | 10.0 (9.8-10.1) | 3.8 (3.7-3.8) | 18.6 (17.7-19.6) | 11.3 (10.0-12.9) | 65.2 (56.8-74.4) | 24.2 (21.5-27.1) |
| Slovenia | 2,078,932 | 90.8 (88.2-93.4) | 28.6 (27.8-29.0) | 8.0 (7.9-8.0) | 11.6 (11.4-11.7) | 4.7 (4.6-4.7) | 24.3 (23.1-25.5) | 12.3 (11.0-13.9) | 159.2 (137.3-184.8) | 49.6 (44.6-54.9) |
| Country | Thousands | N (95% CI) | N (95% CI) | N (95% CI) | N (95% CI) | N (95% CI) | N (95% CI) | N (95% CI) | N (95% CI) | N (95% CI) |
| Spain | 46,754,783 | 91.9 (90.5-93.2) | 27.9 (27.2-28.3) | 8.0 (7.9-8.1) | 11.4 (11.2-11.5) | 4.7 (4.6-4.7) | 25.1 (23.9-26.3) | 12.4 (11.3-13.9) | 100.4 (89.5-112.3) | 37.7 (34.5-41.2) |
| Sweden | 10,099,270 | 95.5 (93.4-97.2) | 27.7 (27.0-28.1) | 7.9 (7.8-8.0) | 11.2 (11.0-11.4) | 4.6 (4.6-4.7) | 24.5 (23.3-25.7) | 12.9 (11.6-14.5) | 90.6 (80.2-102.0) | 35.1 (31.8-38.5) |
| Switzerland | 8,654,618 | 95.6 (92.4-97.8) | 27.4 (26.7-27.9) | 7.7 (7.6-7.7) | 11.0 (10.8-11.2) | 4.5 (4.4-4.5) | 23.5 (22.4-24.7) | 12.9 (11.5-14.6) | 107.1 (93.3-122.5) | 37.8 (33.9-41.9) |
| Tajikistan | 9,537,642 | 51.7 (47.7-55.5) | 13.5 (13.2-13.7) | 2.0 (2.0-2.0) | 3.2 (3.1-3.2) | 1.0 (1.0-1.0) | 3.9 (3.7-4.1) | 7.0 (5.9-8.3) | 4.9 (4.2-5.7) | 1.7 (1.4-2.0) |
| Macedonia | 2,083,380 | 75.1 (72.6-77.5) | 24.5 (23.9-24.9) | 5.9 (5.9-6.0) | 9.0 (8.9-9.2) | 3.3 (3.3-3.4) | 16.0 (15.2-16.8) | 10.2 (9.0-11.6) | 161.4 (138.2-189.8) | 49.8 (44.7-55.3) |
| Turkey | 84,339,067 | 74.4 (70.0-78.4) | 19.1 (18.6-19.4) | 4.1 (4.0-4.1) | 6.2 (6.1-6.3) | 2.2 (2.2-2.2) | 10.4 (9.9-11.0) | 10.1 (8.7-11.7) | 46.4 (40.0-53.7) | 17.8 (15.5-20.2) |
| Turkmenistan | 6,031,187 | 61.6 (58.7-64.8) | 15.3 (15.0-15.6) | 2.6 (2.6-2.6) | 4.1 (4.1-4.2) | 1.3 (1.3-1.3) | 5.6 (5.3-5.9) | 8.3 (7.3-9.7) | N/A<sup>e</sup> | N/A<sup>e</sup> |
| Ukraine | 43,733,759 | 74.6 (68.3-79.8) | 25.9 (25.3-26.3) | 6.7 (6.7-6.8) | 10.0 (9.8-10.2) | 3.9 (3.8-3.9) | 19.6 (18.7-20.6) | 10.1 (8.5-11.9) | 54.1 (45.3-64.7) | 20.8 (17.8-24.3) |
| United Kingdom | 67,886,004 | 90.5 (89.6-91.3) | 26.6 (25.9-27.0) | 7.4 (7.3-7.5) | 10.6 (10.4-10.7) | 4.3 (4.3-4.3) | 22.7 (21.6-23.9) | 12.2 (11.2-13.6) | 101.1 (90.8-112.5) | 37.7 (34.6-40.9) |
| Uzbekistan | 33,469,199 | 62.9 (59.3-66.0) | 15.5 (15.1-15.8) | 2.6 (2.6-2.7) | 4.2 (4.2-4.3) | 1.3 (1.3-1.3) | 5.6 (5.3-5.9) | 8.5 (7.4-9.8) | 6.1 (5.3-7.0) | 2.5 (2.2-2.9) |
| **EURO** | **930,700,857** | **82.6 (79.4-85.7)** | **25.3 (24.7-25.8)** | **6.7 (6.7-6.8)** | **9.8 (9.7-10.0)** | **3.9 (3.9-3.9)** | **20.2 (19.2-21.2)** | **11.2 (9.9-12.8)** | **67.4 (58.8-77.1)** | **25.5 (22.8-28.4)** |

Bosnia & Herz, Bosnia and Herzegovina. CI, credible interval. HAQ, Healthcare Access and Quality. Macedonia, Republic of North Macedonia. N, number. Rep Moldova, Republic of Moldova. Russia, Russian Federation.
<sup>a</sup>The population size and the demographic age-structure were extracted from the United Nations World Population Prospects database [1].
<sup>b</sup>HAQ Index was extracted from the Global Burden of Disease study [4].
<sup>c</sup>The estimate for the incidence rate was based on the average of the two estimates generated by the reported COVID-19 deaths method and the reported COVID-19 cases method.
<sup>d</sup>The estimate for the proportion of the population infected was based on the average of the two estimates generated by the reported COVID-19 deaths method and the reported COVID-19 cases method.
<sup>e</sup>No reported COVID-19 cases and deaths.

**Table S5:** Estimates for key SARS-CoV-2 epidemiologic indicators for countries and territories with a population of at least one million [1] in the WHO South-East Asia Region (SEARO). Classification of infection severity and criticality was per WHO infection severity classifications [2]. Classification of COVID-19 death was also per WHO guidelines [3].

| South-East Asia Region | Total population [1]<sup>a</sup> | HAQ Index [4]<sup>b</sup> | Infection acute-care bed hospitalization rate (per 1,000 infections) | Infection ICU bed hospitalization rate (per 1,000 infections) | Infection severity rate (per 1,000 infections) | Infection criticality rate (per 1,000 infections) | Infection fatality rate (per 10,000 infections) | Infection detection rate (%) | Incidence rate since epidemic onset (per 10,000 person-weeks)<sup>c</sup> | Proportion of the population infected (%)<sup>d</sup> |
| --- | --- | --- | --- | --- | --- | --- | --- | --- | --- | --- |
| Country | Thousands | N (95% CI) | N (95% CI) | N (95% CI) | N (95% CI) | N (95% CI) | N (95% CI) | N (95% CI) | N (95% CI) | N (95% CI) |
| Bangladesh | 164,689,383 | 47.6 (44.3-50.9) | 15.6 (15.2-15.9) | 2.9 (2.8-2.9) | 4.4 (4.3-4.4) | 1.5 (1.5-1.5) | 6.6 (6.2-6.9) | 6.4 (5.5-7.6) | 10.2 (8.7-11.9) | 4.3 (3.7-4.9) |
| DPR. Korea | 25,778,815 | 53.4 (49.6-56.9) | 21.3 (20.7-21.6) | 4.7 (4.7-4.8) | 7.3 (7.2-7.4) | 2.6 (2.6-2.6) | 12.1 (11.5-12.7) | 7.2 (6.2-8.5) | N/A<sup>e</sup> | N/A<sup>e</sup> |
| India | 1,380,004,385 | 41.2 (39.1-43.4) | 16.9 (16.5-17.2) | 3.2 (3.2-3.2) | 5.0 (4.9-5.1) | 1.7 (1.7-1.7) | 7.4 (7.0-7.8) | 5.6 (4.9-6.5) | 22.1 (19.0-25.4) | 10.0 (8.7-11.4) |
| Indonesia | 273,523,621 | 44.5 (42.6-46.8) | 17.3 (16.8-17.5) | 3.2 (3.2-3.3) | 5.1 (5.0-5.2) | 1.7 (1.7-1.7) | 7.3 (6.9-7.6) | 6.0 (5.3-7.0) | 11.7 (10.3-13.3) | 5.0 (4.4-5.6) |
| Myanmar | 54,409,794 | 41.6 (38.0-45.5) | 17.1 (16.6-17.3) | 3.1 (3.1-3.2) | 5.0 (4.9-5.1) | 1.6 (1.6-1.6) | 6.9 (6.5-7.3) | 5.6 (4.7-6.8) | 9.3 (7.8-11.0) | 3.7 (3.1-4.3) |
| Nepal | 29,136,808 | 40.0 (36.5-44.4) | 15.7 (15.3-15.9) | 2.9 (2.8-2.9) | 4.4 (4.3-4.4) | 1.5 (1.5-1.5) | 6.5 (6.2-6.9) | 5.4 (4.5-6.6) | 22.8 (18.6-27.6) | 10.4 (8.6-12.3) |
| Sri Lanka | 21,413,250 | 70.6 (66.3-75.3) | 21.6 (21.1-22.0) | 4.9 (4.8-4.9) | 7.4 (7.3-7.6) | 2.7 (2.7-2.7) | 12.5 (11.9-13.2) | 9.6 (8.3-11.2) | 2.8 (2.4-3.2) | 1.3 (1.1-1.5) |
| Thailand | 69,799,978 | 69.5 (66.5-72.6) | 23.7 (23.1-24.1) | 5.7 (5.6-5.7) | 8.6 (8.5-8.8) | 3.2 (3.1-3.2) | 15.3 (14.5-16.0) | 9.4 (8.3-10.8) | 0.1 (0.1-0.2) | 0.1 (0.1-0.1) |
| Timor-Leste | 1,318,442 | 43.4 (37.2-51.9) | 13.9 (13.6-14.1) | 2.3 (2.3-2.4) | 3.5 (3.4-3.5) | 1.2 (1.1-1.2) | 5.1 (4.8-5.3) | 5.9 (4.6-7.7) | 0.1 (0.1-0.1) | 0.0 (0.0-0.0) |
| **SEARO** | **2,020,074,476** | **43.6 (41.3-46.1)** | **17.2 (16.8-17.4)** | **3.3 (3.2-3.3)** | **5.1 (5.0-5.2)** | **1.7 (1.7-1.7)** | **7.6 (7.3-8.0)** | **5.9 (5.1-6.9)** | **18.1 (15.6-20.8)** | **8.1 (7.0-9.2)** |

CI, credible interval. DPR Korea, Democratic People's Republic of Korea. HAQ, Healthcare Access and Quality. N, number.
<sup>a</sup>The population size and the demographic age-structure were extracted from the United Nations World Population Prospects database [1].
<sup>b</sup>HAQ Index was extracted from the Global Burden of Disease study [4].
<sup>c</sup>The estimate for the incidence rate was based on the average of the two estimates generated by the reported COVID-19 deaths method and the reported COVID-19 cases method.
<sup>d</sup>The estimate for the proportion of the population infected was based on the average of the two estimates generated by the reported COVID-19 deaths method and the reported COVID-19 cases method.
<sup>e</sup>No reported COVID-19 cases and deaths.

**Table S6:** Estimates for key SARS-CoV-2 epidemiologic indicators for countries and territories with a population of at least one million [1] in the WHO Western Pacific Region (WPRO). Classification of infection severity and criticality was per WHO infection severity classifications [2]. Classification of COVID-19 death was also per WHO guidelines [3].

| Western Pacific Region | Total population [1]<sup>a</sup> | HAQ Index [4]<sup>b</sup> | Infection acute-care bed hospitalization rate (per 1,000 infections) | Infection ICU bed hospitalization rate (per 1,000 infections) | Infection severity rate (per 1,000 infections) | Infection criticality rate (per 1,000 infections) | Infection fatality rate (per 10,000 infections) | Infection detection rate (%) | Incidence rate since epidemic onset (per 10,000 person-weeks)<sup>c</sup> | Proportion of the population infected (%)<sup>d</sup> |
| --- | --- | --- | --- | --- | --- | --- | --- | --- | --- | --- |
| Country | Thousands | N (95% CI) | N (95% CI) | N (95% CI) | N (95% CI) | N (95% CI) | N (95% CI) | N (95% CI) | N (95% CI) | N (95% CI) |
| Australia | 25,499,881 | 95.9 (94.8-96.8) | 24.7 (24.0-25.1) | 6.5 (6.4-6.6) | 9.4 (9.3-9.6) | 3.7 (3.7-3.8) | 19.3 (18.4-20.3) | 13.0 (11.8-14.4) | 2.9 (2.7-3.1) | 1.4 (1.3-1.5) |
| Cambodia | 16,718,971 | 39.4 (36.4-42.5) | 14.9 (14.6-15.2) | 2.5 (2.5-2.5) | 3.9 (3.9-4.0) | 1.3 (1.3-1.3) | 5.4 (5.1-5.6) | 5.3 (4.5-6.3) | 0.0 (0.0-0.0) | 0.0 (0.0-0.0) |
| China | 1,439,323,774 | 77.9 (76.5-78.9) | 22.8 (22.2-23.1) | 5.1 (5.1-5.1) | 8.0 (7.9-8.1) | 2.8 (2.8-2.8) | 12.9 (12.3-13.6) | 10.5 (9.5-11.8) | 0.3 (0.3-0.3) | 0.1 (0.1-0.2) |
| Hong Kong | 7,496,988 | 77.9 (76.5-78.9) | 27.5 (26.8-28.0) | 7.4 (7.3-7.4) | 10.9 (10.7-11.1) | 4.3 (4.2-4.3) | 21.8 (20.8-22.9) | 10.5 (9.5-11.8) | 2.1 (1.9-2.3) | 0.9 (0.9-1.0) |
| Japan | 126,476,458 | 94.1 (93.5-94.6) | 32.6 (31.7-33.1) | 10.5 (10.4-10.6) | 14.3 (14.1-14.5) | 6.3 (6.2-6.3) | 34.9 (33.3-36.6) | 12.7 (11.7-14.1) | 2.2 (2.0-2.4) | 1.1 (1.0-1.2) |
| Lao PDR | 7,275,556 | 36.6 (32.6-41.1) | 14.3 (14.0-14.6) | 2.3 (2.3-2.4) | 3.6 (3.6-3.7) | 1.2 (1.1-1.2) | 4.9 (4.6-5.1) | 5.0 (4.1-6.1) | 0.0 (0.0-0.0) | 0.0 (0.0-0.0) |
| Malaysia | 32,365,998 | 68.1 (65.9-70.2) | 17.6 (17.2-17.9) | 3.4 (3.4-3.5) | 5.4 (5.3-5.5) | 1.8 (1.8-1.8) | 8.2 (7.8-8.6) | 9.2 (8.2-10.5) | 5.2 (4.6-5.9) | 2.5 (2.2-2.8) |
| Mongolia | 3,278,292 | 53.4 (49.1-57.6) | 15.3 (14.9-15.5) | 2.6 (2.5-2.6) | 4.1 (4.0-4.1) | 1.3 (1.3-1.3) | 5.2 (4.9-5.5) | 7.2 (6.1-8.6) | 0.6 (0.5-0.7) | 0.3 (0.2-0.3) |
| New Zealand | 4,822,233 | 92.4 (90.3-94.3) | 25.1 (24.5-25.5) | 6.6 (6.5-6.7) | 9.6 (9.5-9.8) | 3.8 (3.8-3.8) | 19.3 (18.4-20.3) | 12.5 (11.3-14.1) | 0.7 (0.6-0.7) | 0.3 (0.3-0.3) |
| Pap. New Gui | 8,947,027 | 31.8 (26.2-37.4) | 13.8 (13.4-14.0) | 2.1 (2.1-2.1) | 3.3 (3.2-3.4) | 1.0 (1.0-1.0) | 4.0 (3.8-4.2) | 4.3 (3.3-5.6) | 0.4 (0.3-0.5) | 0.1 (0.1-0.2) |
| Philippines | 109,581,085 | 51.2 (47.9-54.4) | 15.7 (15.3-16.0) | 2.8 (2.8-2.8) | 4.4 (4.3-4.4) | 1.4 (1.4-1.5) | 6.3 (6.0-6.7) | 6.9 (6.0-8.1) | 14.8 (12.8-17.1) | 6.9 (5.9-7.9) |
| Rep Korea | 51,269,183 | 90.3 (85.6-93.9) | 26.4 (25.8-26.9) | 6.7 (6.7-6.8) | 10.1 (10.0-10.3) | 3.8 (3.8-3.9) | 18.9 (18.0-19.9) | 12.2 (10.7-14.0) | 1.9 (1.7-2.2) | 0.9 (0.8-1.1) |
| Taiwan | 23,816,775 | 85.4 (82.5-88.2) | 25.8 (25.2-26.2) | 6.5 (6.4-6.5) | 9.8 (9.7-10.0) | 3.7 (3.7-3.7) | 18.2 (17.4-19.2) | 11.6 (10.3-13.2) | 0.0 (0.0-0.1) | 0.0 (0.0-0.0) |
| Singapore | 5,850,343 | 90.6 (87.2-93.3) | 24.8 (24.2-25.2) | 5.7 (5.6-5.7) | 9.0 (8.9-9.2) | 3.2 (3.1-3.2) | 14.6 (13.9-15.4) | 12.3 (10.9-13.9) | 9.0 (7.9-10.2) | 4.3 (3.7-4.8) |
| Viet Nam | 97,338,583 | 60.3 (56.3-64.1) | 18.4 (18.0-18.7) | 3.8 (3.7-3.8) | 5.8 (5.8-5.9) | 2.0 (2.0-2.0) | 9.5 (9.0-10.0) | 8.2 (7.0-9.6) | 0.0 (0.0-0.1) | 0.0 (0.0-0.0) |
| **WPRO** | **1,960,061,147** | **76.4 (74.7-77.8)** | **22.8 (22.2-23.1)** | **5.3 (5.2-5.3)** | **8.1 (8.0-8.2)** | **2.9 (2.9-2.9)** | **13.9 (13.2-14.6)** | **10.3 (9.3-11.6)** | **1.4 (1.2-1.6)** | **0.7 (0.6-0.7)** |

CI, credible interval. HAQ, Healthcare Access and Quality. Lao PDR, Lao People's Democratic Republic. N, number. Pap New Gui, Papua New Guinea. Rep Korea, Republic of Korea.
<sup>a</sup>The population size and the demographic age-structure were extracted from the United Nations World Population Prospects database [1].
<sup>b</sup>HAQ Index was extracted from the Global Burden of Disease study [4].
<sup>c</sup>The estimate for the incidence rate was based on the average of the two estimates generated by the reported COVID-19 deaths method and the reported COVID-19 cases method.
<sup>d</sup>The estimate for the proportion of the population infected was based on the average of the two estimates generated by the reported COVID-19 deaths method and the reported COVID-19 cases method.

## References

1. John Hopkins University Coronavirus resource center. COVID-19 Dashboard by the Center for Systems Science and Engineering (CSSE) at Johns Hopkins University (JHU). Available from: https://coronavirus.jhu.edu/map.html. Accessed on July 24, 2020.

2. Anand, S., et al., Prevalence of SARS-CoV-2 antibodies in a large nationwide sample of patients on dialysis in the USA: a cross-sectional study. Lancet, 2020.

3. Havers, F.P., et al., Seroprevalence of Antibodies to SARS-CoV-2 in 10 Sites in the United States, March 23-May 12, 2020. JAMA Intern Med, 2020.

4. Wu, S.L., et al., Substantial underestimation of SARS-CoV-2 infection in the United States. Nat Commun, 2020. 11(1): p. 4507.

5. Stringhini, S., et al., Seroprevalence of anti-SARS-CoV-2 IgG antibodies in Geneva, Switzerland (SEROCoV-POP): a population-based study. Lancet, 2020. 396(10247): p. 313–319.

6. Ioannidis, J.P., The infection fatality rate of COVID-19 inferred from seroprevalence data. Available at: https://www.medrxiv.org/content/10.1101/2020.05.13.20101253v1.full.pdf. Last accessed July 2, 2020. 2020.

7. Lumley, S.F., et al., Antibody Status and Incidence of SARS-CoV-2 Infection in Health Care Workers. N Engl J Med, 2020.

8. Al-Thani, M.H., et al., Seroprevalence of SARS-CoV-2 infection in the craft and manual worker population of Qatar. medRxiv, 2020: p. 2020.11.24.20237719 (non-peer-reviewed preprint).

9. Coyle, P.V., et al., SARS-CoV-2 seroprevalence in the urban population of Qatar: An analysis of antibody testing on a sample of 112,941 individuals. medRxiv, 2021: p. 2021.01.05.21249247.

10. Jeremijenko, A., et al., Evidence for and level of herd immunity against SARS-CoV-2 infection: the ten-community study. medRxiv, 2020: p. 2020.09.24.20200543 (non-peer-reviewed preprint).

11. Ayoub, H.H., et al., Mathematical modeling of the SARS-CoV-2 epidemic in Qatar and its impact on the national response to COVID-19. in press. URL: https://www.medrxiv.org/content/medrxiv/early/2020/11/10/2020.11.08.20184663.full.pdf, 2021.

12. Seedat, S., et al., SARS-CoV-2 infection hospitalization, severity, criticality, and fatality rates. medRxiv 2020.11.29.20240416 (non-peer-reviewed preprint), 2020.

13. World Health Organization, Clinical management of COVID-19. Available from: https://www.who.int/publications-detail/clinical-management-of-covid-19. Accessed on: May 31st 2020. 2020.

14. World Health Organization, International guidelines for certification and classification (coding) of COVID-19 as cause of death. Available from: https://www.who.int/classifications/icd/Guidelines_Cause_of_Death_COVID-19-20200420-EN.pdf?ua=1. Document Number: WHO/HQ/DDI/DNA/CAT. Accessed on June 1, 2020.. 2020.

15. Abu-Raddad, L.J., et al., Characterizing the Qatar advanced-phase SARS-CoV-2 epidemic. medRxiv, 2020: p. 2020.07.16.20155317v2 (non-peer-reviewed preprint).

16. Abu-Raddad, L.J., et al., Two prolonged viremic SARS-CoV-2 infections with conserved viral genome for two months. Infect Genet Evol, 2020. 88: p. 104684.

17. Abu-Raddad, L.J., et al., Assessment of the risk of SARS-CoV-2 reinfection in an intense re-exposure setting. Clinical Infectious Diseases, 2020. ciaa1846. doi: 10.1093/cid/ciaa1846..

18. Al Kuwari, H.M., et al., Epidemiological investigation of the first 5685 cases of SARS-CoV-2 infection in Qatar, 28 February–18 April 2020. BMJ Open, 2020. 10(10): p. e040428.

19. Ayoub, H.H., et al., Epidemiological impact of prioritizing SARS-CoV-2 vaccination by antibody status: Mathematical modeling analyses. medRxiv, 2021: p. 2021.01.10.21249382.

20. Nasrallah, G.K., et al., Are commercial antibody assays substantially underestimating SARS-CoV-2 ever infection? An analysis on a population-based sample in a high exposure setting. medRxiv, 2020: p. 2020.12.14.20248163.

21. Abu-Raddad, L.J., et al., SARS-CoV-2 reinfection in a cohort of 43,000 antibody-positive individuals followed for up to 35 weeks. medRxiv, 2021: p. 2021.01.15.21249731.

22. Ayoub, H.H., et al., Characterizing key attributes of the epidemiology of COVID-19 in China: Model-based estimations. Global Epidemiology, 2020. 100042.

23. Ayoub, H.H., et al., Age could be driving variable SARS-CoV-2 epidemic trajectories worldwide. Plos One, 2020. 15(8).

24. Davies, N.G., et al., Age-dependent effects in the transmission and control of COVID-19 epidemics. Nat Med, 2020.

25. Mizumoto, K., R. Omori, and H. Nishiura, Age specificity of cases and attack rate of novel coronavirus disease (COVID-19). medRxiv, 2020: p. 2020.03.09.20033142.

26. United Nations Department of Economic and Social Affairs Population Dynamics, The 2019 Revision of World Population Prospects. Available from https://population.un.org/wpp/. Accessed on March 1st, 2020. 2020.

27. WHO Coronavirus Disease (COVID-19) Dashboard. Available from: https://covid19.who.int/. Accessed on December 31, 2020. 2020.

28. Fullman, N., et al., Measuring performance on the Healthcare Access and Quality Index for 195 countries and territories and selected subnational locations: a systematic analysis from the Global Burden of Disease Study 2016. The Lancet, 2018. 391(10136): p. 2236–2271.

29. Faes, C., et al., Time between Symptom Onset, Hospitalisation and Recovery or Death: Statistical Analysis of Belgian COVID-19 Patients. Int J Environ Res Public Health, 2020. 17(20).

30. COVID-19 Clinical Information Network (CO-CIN), CO-CIN: COVID-19 -Time from symptom onset until death in UK hospitalised patients, 7 October 2020. Available at: https://www.gov.uk/government/publications/co-cin-covid-19-time-from-symptom-onset-until-death-in-uk-hospitalised-patients-7-october-2020. Accessed January 20, 2021.

31. Davies, N.G., et al., Association of tiered restrictions and a second lockdown with COVID-19 deaths and hospital admissions in England: a modelling study. Lancet Infect Dis, 2020.

32. Centers for Disease Control and Prevention, COVID-19 Pandemic Planning Scenarios - September 10, 2020. Available at: https://www.cdc.gov/coronavirus/2019-ncov/hcp/planning-scenarios.html#table-2. Accessed January 20, 2021.

33. World Health Organization, World Health Organization Countries Classification. Available at: https://www.who.int/countries. Accessed January 22, 2021.

34. MATLAB®, The Language of Technical Computing. The MathWorks, Inc. 2019.

35. Anderson, R.M., et al., How will country-based mitigation measures influence the course of the COVID-19 epidemic? Lancet, 2020. 395(10228): p. 931–934.

36. Britton, T., F. Ball, and P. Trapman, A mathematical model reveals the influence of population heterogeneity on herd immunity to SARS-CoV-2. Science, 2020. 369(6505): p. 846–849.

37. Lauer, S.A., et al., The Incubation Period of Coronavirus Disease 2019 (COVID-19) From Publicly Reported Confirmed Cases: Estimation and Application. Ann Intern Med, 2020. 172(9): p. 577–582.

38. Wu, Z. and J.M. McGoogan, Characteristics of and Important Lessons From the Coronavirus Disease 2019 (COVID-19) Outbreak in China: Summary of a Report of 72314 Cases From the Chinese Center for Disease Control and Prevention. JAMA, 2020.

39. Ghina R Mumtaz, et al., Can the COVID-19 pandemic still be suppressed? Putting essential pieces togetherJ. Journal of Global Health Reports, 2020. 4.

40. Meyerowitz-Katz, G. and L. Merone, A systematic review and meta-analysis of published research data on COVID-19 infection-fatality rates. Int J Infect Dis, 2020.

41. Hauser, A., et al., Estimation of SARS-CoV-2 mortality during the early stages of an epidemic: A modeling study in Hubei, China, and six regions in Europe. PLoS Med, 2020. 17(7): p. e1003189.

42. Salje, H., et al., Estimating the burden of SARS-CoV-2 in France. Science, 2020.

43. R. Verity, et al., Estimates of the severity of coronavirus disease 2019: a model-based analysis. The Lancet, 2020. 20: p. 30243–7.

44. Brazeau N. F., et al., Report 34: COVID-19 Infection Fatality Ratio: Estimates from Seroprevalence. Imperial College London (29-10-2020), doi https://doi.org/10.25561/83545. 2020.

45. Tableau 2017: Seattle, Washington. p. Online: https://www.tableau.com/.

## References

1. United Nations Department of Economic and Social Affairs Population Dynamics, The 2019 Revision of World Population Prospects. Available from https://population.un.org/wpp/. Accessed on March 1st, 2020. 2020.

2. World Health Organization, Clinical management of COVID-19. Available from: https://www.who.int/publications-detail/clinical-management-of-covid-19. Accessed on: May 31st 2020. 2020.

3. World Health Organization, International guidelines for certification and classification (coding) of COVID-19 as cause of death. Available from: https://www.who.int/classifications/icd/Guidelines_Cause_of_Death_COVID-19-20200420-EN.pdf?ua=1. Document Number: WHO/HQ/DDI/DNA/CAT. Accessed on June 1, 2020.. 2020.

4. Fullman, N., et al., Measuring performance on the Healthcare Access and Quality Index for 195 countries and territories and selected subnational locations: a systematic analysis from the Global Burden of Disease Study 2016. The Lancet, 2018. 391(10136): p. 2236–2271.

